# Multiplexed temporal SWCNT biosensor combined with convolutional autoencoding identifies ALS-associated serum protein corona signatures

**DOI:** 10.64898/2026.06.08.26354966

**Authors:** Riccardo Sirtori, Rodrigo Monroy Lopez, Huifang Li, Chang Liu, Nicholas Fisk, Daniel Roxbury, Claudia Fallini

## Abstract

Amyotrophic lateral sclerosis (ALS) lacks a validated blood-based diagnostic, and the field is increasingly moving from single-molecule markers toward integrative, multi-component signatures. Here we present a liquid-biopsy strategy that transduces disease-dependent serum–nanoparticle interactions into a learnable near-infrared spectral phenotype. A sensor array of twelve DNA-functionalized single-walled carbon nanotube (SWCNT) chiralities, functionalized with (GT)_6_ ssDNA coupled with a deep learning model was tested on serum from 20 ALS patients and 19 sex-matched controls (n = 39, TargetALS). Our multiplexed sensor design (12 SWCNT chiralities) and data acquisition strategy based on excitation–emission matrices acquired at three timepoints (0, 6, 24 h) was conceived to maximize sensor carried information. Indeed, we show that the array generates partially independent temporal dynamics across chiralities governed primarily by tube diameter. To decode this multiplexed, time-resolved signal, we trained a dual-objective convolutional autoencoder that jointly optimizes reconstruction and classification, achieving 84.6% cross-validated accuracy (AUC = 0.87).

Selected latent features were reproducible across an independent same-subject experimental batch and correlated with serum neurofilament light chain, linking the spectral phenotype to a clinically relevant neurodegeneration marker. Mass spectrometry supported a molecular basis for discrimination, revealing an ALS-biased protein corona enriched in adaptive-immune and inflammatory proteins. Together, these results establish proof of principle that time-resolved, multi-chirality SWCNT spectral sensing can compress complex serum composition into a reproducible near-infrared biomarker signature for ALS.

## 1. Introduction

Amyotrophic lateral sclerosis (ALS) is a rapidly progressive neurodegenerative disease mainly affecting people between 60 and 70 years old, with a median survival of 2–5 years^1^. Estimates suggest that the progressive aging of the world population will lead to a dramatic increase in the number of ALS cases in the next decade^2^, highlighting the urgent need for novel diagnostic and therapeutic tools. An early diagnosis is essential for providing ALS patients with therapeutic options that can help slow disease progression and preserve quality of life^3^. However, the absence of clinically validated biological or biochemical markers makes the diagnostic process, which relies on clinical symptoms and the exclusion of similar pathologies, extremely lengthy, with average diagnostic delays of approximately 12 months from symptom onset. This significantly reduces the impact of potential treatments, whose maximum efficacy in animal models is achieved at a pre-symptomatic stage^3^.

Current biomarkers under pre-clinical development for ALS rely on detecting a single molecule in blood or cerebrospinal fluid (CSF). Increased levels of neurofilament light chain (NfL) show high sensitivity for ALS and good correlation with disease progression but poor specificity^4^.

Detection of total or phosphorylated TDP-43 in CSF, a hallmark of ALS which is mechanistically connected to disease processes, is inconsistent during the disease course and displays low detectability^5^. Recent efforts have focused on cryptic peptides, downstream products of TDP-43 loss of splicing regulatory function, as promising biomarkers; however, only one such peptide, HDGFL2, has been identified so far^6^. While replication studies are ongoing, issues with developing high-sensitivity assays requiring custom antibodies have emerged^7^. These observations highlight the need for a novel approach that integrates multiple signals into a specific and sensitive biomarker.

Blood biosensors represent an emerging technology for developing integrative biomarkers for multifactorial diseases like ALS, whose complex nature may not be reducible to a single or small group of biomolecules. This is particularly relevant for sporadic ALS (sALS), which accounts for approximately 90% of all cases^1^, where no genetic biomarker exists and the pathologic cascade is still largely unknown. Single-walled carbon nanotubes (SWCNTs) offer considerable promise in this space, as they can detect and integrate many biological features in a single biosensor. SWCNTs are graphene cylinders exhibiting near-infrared (NIR) fluorescence that is highly sensitive to their biochemical environment. Upon exposure to biological samples, the formation of a protein corona (PC) - a nanoscale coating of proteins with high affinity for the material surface - modulates SWCNT NIR optical features (i.e., excitation peak wavelength, emission peak wavelength, and intensity), generating a unique spectral fingerprint reflective of the sample’s molecular composition^8,9^. This approach, combined with machine learning, has demonstrated both high diagnostic performance (e.g., 87% sensitivity and 98% specificity for ovarian cancer detection from serum)^10^, and the ability to detect disease-specific PC signatures, as demonstrated for Alzheimer’s disease using patients’ plasma^11^.

Here, we present a novel approach combining SWCNT NIR fluorescence spectral fingerprints with deep machine learning to develop and characterize an integrative multicomponent biomarker (MCB)^12^ for detecting ALS disease state from patients’ blood serum. Our approach rests on two pillars: (i) full exploitation of the molecular profiling capacity of SWCNTs through multi-chirality time-resolved data acquisition, and (ii) data-driven convolutional autoencoder for unbiased feature extraction. This approach is innovative as it leverages two well-established properties of SWCNTs simultaneously: the Vroman effect and chirality selectivity. The former describes the adsorption kinetics of proteins on nanoparticle surfaces, where high-abundance proteins adsorb first and are gradually replaced by high-affinity proteins over time^13,14^. The latter arises because of the specific way the graphene sheet is rolled (defined by the chiral vector), which confers each SWCNT species distinct physicochemical properties (e.g. varying diameter and underlying carbon lattice orientation) and thus different NIR spectral features and adsorption kinetics^15,16,17^. We demonstrate that a multi-chirality SWCNT system leverages the multiplexed and transient nature of the protein corona to deliver a higher information content than a single-chirality sensor, but requires a more sophisticated analytical framework. For this reason, we designed a convolutional autoencoder that learns discriminative features by jointly optimizing reconstruction and classification losses, enabling data-driven interpretation of complex multi-chirality spectroscopic images. The extracted features were subsequently confirmed as discriminative through a simple linear support vector machine (SVM), validated for robustness against batch effects, and correlated with a known neurodegeneration biomarker (NfL levels).

Finally, the biological basis of the sensor response was validated through mass spectrometry analysis of the protein corona.

## 2. Methods

### 2.1 Patient cohort and sample collection

Blood serum samples were obtained from 20 ALS patients and 19 sex-matched healthy controls (total *n* = 39) from TargetALS. ALS patients were diagnosed according to the revised El Escorial criteria^18^. Disease severity was assessed using the ALS Functional Rating Scale–Revised (ALSFRS-R)^19^, and disease progression rate was quantified as the ALSFRS-R slope (mean = −2.60 ± 3.86 points/month). Serum neurofilament light chain (NfL) concentration measured by single-molecule array (Simoa^20^) was provided by TargetALS Informatics Core (Supp. Table 1). Upon receipt, serum samples were aliquoted and stored at −80 °C until use.

**Table 1.**
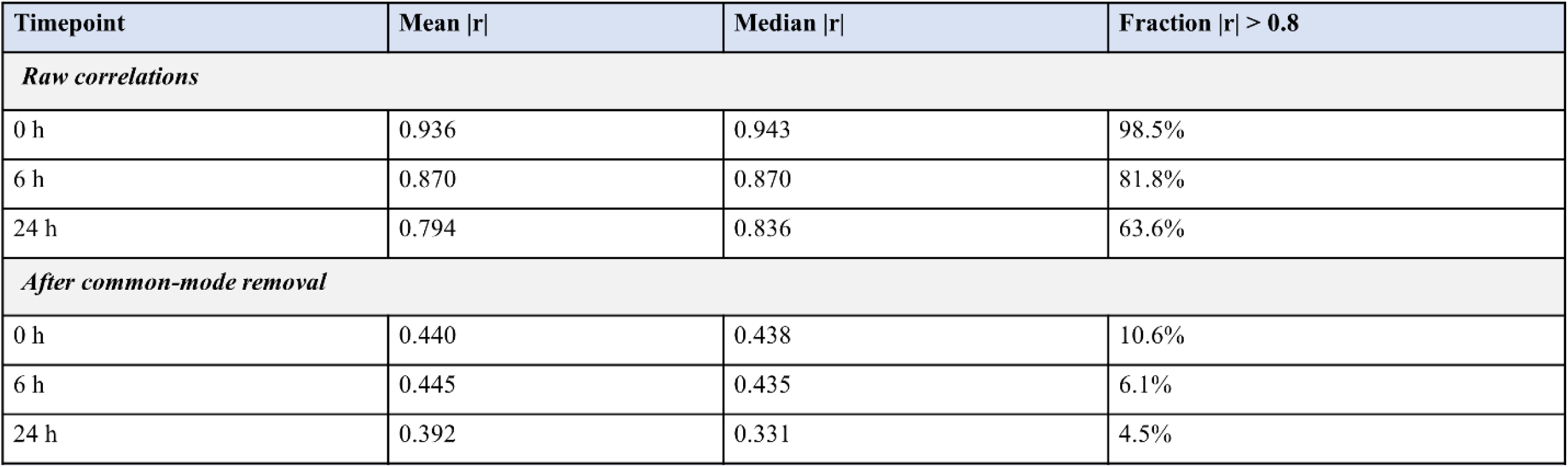
Inter-chirality correlation metrics by timepoint, before and after common-mode removal. Mean |*r*| = mean absolute Pearson correlation across all 66 off-diagonal chirality pairs. Common-mode removal subtracts the per-observation mean across chiralities to isolate chirality-specific dynamics from shared intensity drift.

| Timepoint | Mean $\vert r\vert $ | Median $\vert r\vert $ | Fraction $\vert r\vert > 0.8$ |
| --- | --- | --- | --- |
| *Raw correlations* |  |  |  |
| 0 h | 0.936 | 0.943 | 98.5% |
| 6 h | 0.870 | 0.870 | 81.8% |
| 24 h | 0.794 | 0.836 | 63.6% |
| *After common-mode removal* |  |  |  |
| 0 h | 0.440 | 0.438 | 10.6% |
| 6 h | 0.445 | 0.435 | 6.1% |
| 24 h | 0.392 | 0.331 | 4.5% |

Before use, serum samples are processed using Amicon Ultra Centrifugal filters 3 kDa MWCO to standardize the buffer composition and remove low-molecular-weight molecules that could influence protein corona formation.

### 2.2 SWCNT sensor preparation and NIR spectroscopy

For ssDNA-SWCNT sensor production, we mixed multi-chirality HiPco SWCNT powder (Nanointegris) with ssDNA (Integrated DNA Technologies, Inc) in 0.1 M NaCl at fixed mass ratio (mass SWCNT:mass ssDNA, 1:5) and 1 mL total volume. The sample was ultrasonicated using a 1/8″ tapered microtip for 30 min at 40 % amplitude in an ice bath (Sonics Vibracell VCX-130; Sonics and Materials). The resulting suspensions were ultra-centrifuged (Beckman Optima MAX-XP) for 30 min at 250,000 g and 4 °C, and the top ∼80 % of the supernatant was collected. Concentrations were determined using a UV/vis/NIR spectrophotometer (JASCO, Tokyo, Japan) and an extinction coefficient of A910 = 0.02554 L mg^−1^ cm^−1^ ^21^.

SWCNT incubation in serum samples was performed at the final concentration of 1 mg L^-1^. Serum samples (diluted in PBS) were mixed with the SWCNT sensor suspension, and excitation–emission matrices (EEMs) were acquired using NS super NanoSpectralyzer (Applied NanoFluorescence) at three timepoints: 0 h (baseline, immediately after serum application), 6 h, and 24 h post-exposure. EEMs were recorded across excitation wavelengths of approximately 500–850 nm and emission wavelengths of approximately 850–1700 nm. Each EEM was stored as the mean of three two-dimensional matrices before analysis. Two independent experimental batches were collected: Batch 1 (*n* = 39) for model development, and Batch 2 (*n* = 39, same subjects) for batch-transfer validation.

### 2.3 Mass spectrometry of the protein corona

To characterize the protein corona formed on SWCNTs, parallel experiments were performed with pooled ALS (*n* = 3) and control (*n* = 3) serum samples. After 24 h incubation, SWCNT-PC complexes underwent 3 cycles of washing and spin pull-down steps to discard unbound proteins. Bound proteins were then eluted from SWCNTs at 60 ° C for 1 hour in DTT-urea buffer (5 mM DTT, 8 M urea, 50 mM Tris-HCl pH 8). Before alkylation buffer exchange was carried out using Amicon Ultra Centrifugal filters 3 kDa MWCO (Millipore). The protein extracts were then alkylated in 15 mM iodoacetamide for 30 minutes at room temperature, followed by quenching with 500 mM DTT (10 mM). Trypsin enzymatic digestion was performed in 50 mM Tris-HCl (pH 8) for 12 hours at 37 ° C, and peptides were purified using Amicon Ultra Centrifugal filters 30 kDa MWCO to remove any leftover SWCNT. Peptides were then desalted using C18 Pierce purification columns (Thermo Fisher Scientific) and resuspended in 0.1 % formic acid (Thermo Fisher Scientific). Instrumental data acquisition was performed using an Orbitrap Exploris 240 mass spectrometer coupled to a Vanquish Flex UHPLC system (Thermo Fisher Scientific, USA). Data-independent acquisition (DIA) was conducted over a 120 min run, including a 5 min equilibration period, at a flow rate of 0.050 mL/min. Peptide separation was achieved using an Acquity UPLC Peptide BEH C18 column (2.1 × 150 mm, 300 Å, 1.7 μm) equipped with a VanGuard pre-column (2.1 × 5 mm, 300 Å, 1.7 μm). The column temperature was maintained at 50 °C, and the autosampler was set to 10 °C. Mobile phase A consisted of water with 0.1% formic acid, and mobile phase B consisted of acetonitrile with 0.1% formic acid. The gradient was programmed as follows: 2% B at initial conditions, increased to 30% B at 90 min, 50% B at 101 min, and 95% B at 102 min; held at 95% B until 112 min, followed by re-equilibration to 2% B at 114 min. The mass spectrometer was operated in positive ion mode. The DIA method consisted of a full MS scan (m/z 400–1100) acquired at 60,000 resolution, followed by MS/MS acquisition using 69 variable isolation windows (nominal width of 10 m/z with 1 m/z overlap) spanning the same precursor range. Fragmentation was performed using higher-energy collisional dissociation (HCD) at 30% normalized collision energy, and MS/MS spectra were acquired at 60,000 resolution over an m/z range of 145–1450.

DirectDIA analysis was performed using Spectronaut (version 17) as previously described^22,23^. A human proteome FASTA database (UniProt ID UP000005640)^24^ was used for library generation. Default settings in Spectronaut were applied unless otherwise noted. Trypsin/P was specified as the protease, with carbamidomethylation set as a fixed modification. Variable modifications included protein N-terminal acetylation and methionine oxidation. The false discovery rate (FDR) was controlled at 1% at the peptide, protein, and peptide-spectrum match (PSM) levels.

This statistical control, in combination with mProphet scoring and IDPicker algorithms, was used to minimize false identifications. Extracted ion chromatogram (XIC) retention time alignment was performed using a dynamic window with tolerances automatically optimized by Spectronaut, while other parameters were maintained at default settings. Protein intensities were normalized in Spectronaut (version 17) using the local normalization strategy.

Prior to statistical testing, likely epithelial/skin contaminants were removed based on protein annotation, including entries containing terms such as keratin, filaggrin, hornerin, desmoglein, desmocollin, corneodesmosin, cornulin, involucrin, loricrin, trichohyalin, late cornified envelope, small proline-rich, epidermal, cuticular, and related descriptors. Raw intensities were log2-transformed and median-normalized across samples. Missing values were then imputed on the normalized log2 scale using a left-shifted Gaussian distribution, with imputed values drawn from a normal distribution centered at the observed sample mean minus 1.8 standard deviations and with a width of 0.3 standard deviations, consistent with the assumption that missing values predominantly reflect low-abundance proteins. Differential abundance between ALS and control samples was assessed using Welch’s *t*-test, with Benjamini–Hochberg false discovery rate (FDR) correction for multiple testing^25^.

Physicochemical analyses were performed at the protein level by integrating proteomic outputs with sequence- and structure-derived descriptors. Differentially abundant proteins were linked to physicochemical descriptors to test whether proteins enriched in different corona conditions shared common adsorption-related properties. For sequence-based analyses, protein-group accessions were matched to UniProt entries^24^, and one representative sequence was assigned to each detected protein group. From the amino-acid sequence, glycine fraction, leucine fraction, GRAVY (grand average of hydropathy), and instability index were calculated using BioPython ProtParam/ProteinAnalysis^26^. Glycine and leucine fractions were defined as the proportion of glycine or leucine residues relative to total sequence length. GRAVY was computed as the mean residue hydropathy across the full sequence, and instability index was calculated from dipeptide composition as a sequence-based estimate of intrinsic protein instability. For structure-based analyses, AlphaFold^27,28^ or annotation-derived descriptors were incorporated including mean pLDDT, the fractions of residues with pLDDT < 50 and pLDDT < 70, and predicted helix, sheet, and coil fractions. These variables were used as complementary descriptors of structural order, disorder propensity, and secondary-structure composition.

To evaluate whether ALS- and CTRL-biased corona proteins differed in adsorption-related chemistry, two complementary approaches were used. First, proteins were grouped according to the direction of differential abundance, for example ALS-higher versus CTRL-higher, and the distributions of individual descriptors were compared between these subsets. Second, the continuous log2 fold change value for each protein was used to test whether any descriptor covaried with the degree and direction of enrichment in the corona. Univariate comparisons were performed on a per-feature basis, and p-values were adjusted for multiple testing using the Benjamini–Hochberg false discovery rate procedure^25^. Multivariate analyses were additionally performed on the descriptor matrix to test whether combinations of sequence- and structure-derived properties distinguished ALS-enriched from CTRL-enriched proteins. These analyses therefore assessed whether proteins preferentially represented in one corona state exhibited a distinct adsorption-relevant physicochemical profile.

A separate global selectivity analysis was used to determine whether the (GT)_6_-SWCNT sensor captured a chemically non-random subset of the serum proteome. Proteins identified in the corona dataset were treated as the detected set and compared with the broader serum-proteome reference background assembled from the uploaded serum protein resources. Descriptor distributions were then compared between the detected and background sets to test whether the sensor preferentially enriched proteins with specific physicochemical characteristics.

Accordingly, the ALS-versus-CTRL chemistry analysis was based on protein-level differential abundance metrics, whereas the serum-background selectivity analysis was based solely on protein set membership independent of abundance magnitude.

### 2.4 Chirality-Specific Feature Extraction from EEM Spectra

Physical descriptors were extracted from excitation-emission matrix (EEM) spectroscopic data for each of the 12 SWCNT chiralities at three incubation timepoints (0 h, 6 h, 24 h) using a precision interpolation and curve-fitting pipeline.

For each sample and timepoint, the raw EEM intensity matrix was first upsampled by a factor of 10× along both the emission and excitation axes using bicubic spline interpolation (scipy.interpolate.RectBivariateSpline, kx = ky = 3), transforming the discrete spectral grid into a continuous intensity surface with sub-nanometer resolution. For each chirality, the expected peak position was defined by its known emission and excitation coordinates derived from the DNA-wrapped SWCNT reference chart^15,16^. A local maximum search was then performed within a 2 nm radius of the expected position on the interpolated surface to accommodate minor sample-to-sample shifts in peak location.

A one-dimensional emission intensity profile was extracted at the excitation wavelength of the identified local maximum. To define a physically meaningful fitting window, a gradient-consistency algorithm was applied: starting from the peak, the profile was traversed in both directions along the emission axis, tracking monotonic intensity descent. The walk terminated after two consecutive gradient reversals, preventing the inclusion of noise or neighboring peak contributions. The resulting window was further capped at 2.5× the measured half-width on each side to exclude non-Gaussian tails.

An asymmetric (split) Gaussian function with a constant baseline was fitted to the selected emission profile points. The split Gaussian employed independent width parameters (σ_left, σ_right) for the short- and long-wavelength sides of each peak, capturing the characteristic asymmetry of SWCNT fluorescence emission. Prior to fitting, intensities were normalized to the [0, 1] interval for numerical stability, with parameters subsequently rescaled to physical units.

Seven descriptors were extracted per chirality per timepoint: fitted peak amplitude, emission center wavelength, excitation center wavelength, full width at half maximum (FWHM = 1.177 × (σ_left + σ_right)), area under the curve, and intensity-weighted skewness and kurtosis of the emission profile. Goodness of fit was assessed via R² and RMSE in original intensity units.

When curve fitting did not converge, empirical estimates based on direct half-maximum interpolation and trapezoidal integration were used as fallbacks.

In addition to single-chirality descriptors, pairwise relational features were computed for all 66 chirality pairs: intensity ratios (peak amplitude of chirality A divided by chirality B) and Euclidean distances between peak positions in the emission-excitation plane.

### 2.5 Chirality kinetic independence analysis

To validate the multiplexing capacity of the multi-chirality sensor, we quantified the degree of independence in spectral dynamics across chiralities. For each sample and timepoint, each chirality’s emission peak intensity was calculated as shown in 2.4. Pairwise Pearson correlations were computed between all 66 chirality pairs at each timepoint. Redundancy was quantified as the fraction of pairs with |*r*| > 0.8 and the mean absolute off-diagonal correlation. To isolate chirality-specific dynamics from common-mode intensity drift (shared environmental changes affecting all chiralities similarly), we subtracted the per-observation mean across chiralities from each feature vector. The resulting common-mode-removed correlation matrices were assessed using a permutation test (2,000 permutations) to determine whether the observed mean |*r*| was lower than expected by chance.

Temporal dynamics were analyzed through three complementary frameworks. First, a global PERMANOVA^29^ (9,999 permutations, Euclidean distance) tested whether EEM profiles differed significantly across the three timepoints, with pairwise tests and Holm correction. Second, PERMDISP^30^ assessed whether multivariate dispersion changed across timepoints using a Friedman test on distances to group centroids (tests whether the distances from individual observations to their group centroid differ across timepoints). Third, per-chirality temporal changes were quantified using repeated-measures ANOVA with Benjamini–Hochberg FDR^25^ correction across the 12 chiralities, followed by paired post-hoc comparisons. A linear mixed model (LMM) implemented in statsmodels^31^ with time, chirality, and their interaction as fixed effects and subject as a random intercept tested whether temporal trajectories differed across chiralities. Principal component analysis (PCA) was performed on the 12-chirality feature vectors to visualize temporal trajectories, with paired *t*-tests (Holm-corrected) on PC scores.

To investigate the structural determinants of corona formation kinetics, we fitted a second set of LMMs parameterizing chirality through its two physical properties -tube diameter (d) and chiral angle (θ) -rather than treating it as a categorical factor^15–17^. Diameter and chiral angle were computed from the chiral indices (n, m) using standard relations (d = a_C−C_ √(3(n² + nm + m²)) / π, where a_C−C_= 0.142 nm; θ = arctan(√3 m / (2n + m))). Both were z-standardized to enable direct comparison of effect sizes. The full model included time (categorical: 0 h, 6 h, 24 h), standardized diameter, standardized chiral angle, all two-way interactions, and the three-way time × diameter × chiral angle interaction as fixed effects, with subject as a random intercept.

Nested model comparisons were performed using likelihood ratio (LR) tests^32^, dropping each structural term (and all its interactions) to assess its contribution. Two spectral observables derived from Gaussian fits to each chirality’s emission peak were analyzed: peak fluorescence intensity (Gaussian maximum amplitude) and emission center wavelength (Gaussian emission peak position), providing complementary views of corona-induced spectral modulation.

### 2.6 Convolutional autoencoder architecture

We designed a convolutional autoencoder^33^ with a dual-objective loss function to extract discriminative features from the multi-chirality, time-resolved excitation-emission matrices (EEMs). The architecture consists of three main components: (i) a convolutional encoder, (ii) a convolutional decoder, and (iii) an attention-based temporal aggregation and classification module^34^.

The encoder comprises four convolutional blocks, each consisting of a 2D convolution (3 × 3 kernel, stride 1, padding 1), batch normalization^35^, ReLU activation^36^, and 2 × 2 max pooling, with channel progression 32 → 64 → 128 → 256. The final convolutional block is followed by flattening and a fully connected bottleneck layer that maps each single-timepoint EEM to a 512-dimensional latent representation *z_t_*. For each subject, the encoder is applied independently to the three EEMs corresponding to 0 h, 6 h, and 24 h, yielding latent vectors *z*_0ℎ_, *z*_6ℎ_, and *z*_24ℎ_.

The decoder mirrors the encoder and reconstructs each timepoint EEM from its corresponding individual latent vector, preserving timepoint-specific information during reconstruction.

To integrate the three latent snapshots into a subject-level representation, we used a learned attention mechanism^34^. For each timepoint latent vector, a small neural network computes a scalar importance score, and these scores are normalized with a softmax across timepoints to obtain attention weights *α_i_*. The final aggregated latent vector is therefore:

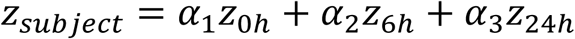

where *α*_1_ + *α*_2_ + *α*_3_ = 1. This attention mechanism allows the model to learn which incubation time-points contribute most strongly to disease discrimination for a given subject.

The aggregated latent vector is then passed to the classification head. In the implemented model, this head is a multilayer perceptron (MLP): it consists of a fully connected layer projecting the latent vector to 128 hidden units, followed by ReLU activation, dropout (*p* = 0.3)^37^, and a final linear layer producing two output values corresponding to the ALS and control classes. These outputs are raw logits; no sigmoid activation is applied within the network, as class probabilities are obtained implicitly through the cross-entropy objective during training.

The model was trained by jointly optimizing reconstruction and classification objectives. Reconstruction loss was defined as the mean squared error (MSE) between each input EEM and its reconstruction, whereas classification loss was defined as cross-entropy loss on the two-class logits. The total loss was:

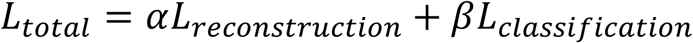

with *α* = 1.0 and *_β_* = 1.0. This dual-objective formulation encourages the shared encoder to learn latent features that are both information-preserving for EEM reconstruction and discriminative for group classification. Training was performed in PyTorch^38^ using the Adam optimizer^39^ with learning rate 1 × 10^-3^ for up to 100 epochs. We trained five independent autoencoder models using stratified 5-fold subject-level cross-validation. All three time-points from the same subject were always assigned to the same fold, preventing information leakage across time-points. Each model was initialized with fresh random weights and trained on a different set of approximately 31 subjects (∼80%), with the remaining ∼8 subjects held out for validation. As a result, the five models share identical architecture and hyperparameters but differ in two ways: (i) each sees a different 80% of the data during training, and (ii) each starts from different random weight initialization, leading to convergence toward different local minima in the loss landscape. This design is motivated by the goal of identifying a robust spectral biomarker: because each model independently discovers discriminative features from a slightly different subset of the cohort, latent dimensions that are consistently informative across multiple models are more likely to reflect genuine biological signal rather than idiosyncratic patterns in a particular training set. The downstream feature selection pipeline (Section 2.8) exploits this multiplicity by pooling all 512 × 5 = 2,560 candidate dimensions and selecting only those that achieve high discriminative performance and remain stable across the LOO folds of the ranking procedure.

### 2.7 Feature extraction and interpretability

To interpret which spectral features drive discrimination, we employed three attribution methods. Gradient saliency computes the absolute gradient of each latent dimension with respect to the input EEM, highlighting pixels whose perturbation most affects the latent representation^40^.

Integrated gradients accumulate gradients along a linear path from a zero-baseline to the actual input, providing a more faithful attribution that satisfies axiomatic completeness^41^. Linear probing trains a linear classifier on each latent dimension independently to predict group membership, with the probe weights indicating which input features are most linearly associated with each latent dimension^42^. An importance score map was constructed by averaging Gradient saliency and Linear probing per pixel normalized values of latent dimensions from each model (pixel importance score = mean[normalize(gradient), normalize(linear)]).

### 2.8 Proxy latent dimension extraction and batch transfer

To create interpretable, batch-robust features from the autoencoder’s learned representations, we developed a two-stage pipeline combining discriminative feature selection with encoder-based knowledge distillation.

**Stage 1: Latent dimension selection.** For each of the 5 cross-validation folds, we computed the attention-weighted latent representation z_agg_ ∈ ℝ^512^ for all 39 subjects, yielding a pool of 2,560 candidate features (512 dimensions × 5 folds). Candidates were ranked using a leak-free AUC procedure: within each leave-one-out (LOO) fold, the univariate AUC for discriminating ALS from controls was computed on the training partition only (*n* = 38), avoiding any information leakage from the held-out sample. The mean AUC across all 39 LOO folds served as the ranking score (|AUC − 0.5|). In addition, a stability metric was computed as the fraction of LOO folds in which a given feature ranked among the top 50; candidates with stability < 0.50 were excluded. Features were then selected iteratively using a greedy forward approach with Spearman redundancy pruning; at each step, the highest-ranked eligible feature was added only if its Spearman |ρ| with all previously selected features remained below 0.85, ensuring complementary information content. After each addition, LOOCV classification performance was evaluated using three classifiers (linear SVM, random forest, and logistic regression) implemented in scikit-learn^43^, and the best AUC across classifiers was tracked. Selection stopped when the monitored AUC failed to improve for more than 2 consecutive steps (patience = 2), and the final feature set was rolled back to the step that achieved peak AUC. The significance of the final feature set was assessed by a permutation test (1000 permutations of the class labels, full LOOCV re-evaluation at each permutation). This procedure yielded 3 non-redundant latent dimensions (dim477 from fold 3, dim103 from fold 1, and dim370 from fold 3, **Supp. Figure 1a**). **Stage 2:** we used the encoder’s frozen weights to analytically reconstruct the full aggregated latent vector with zero free parameters. First, convolutional features at each timepoint are multiplied by the full FC weight matrix and bias to produce per-timepoint latent vectors. Second, the pre-trained attention network (a two-layer neural network with tanh activation) takes these latent snapshots as input and computes a timepoint importance weight for each sample. Third, the attention-weighted average yields z_agg_, from which the target dimension is extracted. Because proxy extraction itself uses frozen pretrained weights and does not fit new parameters to the classification labels, no additional supervised fitting is introduced during proxy construction. However, final classification performance was still interpreted together with permutation testing and batch-transfer validation because latent feature selection was performed on the discovery cohort (**Supp. Figure 1b**).

Batch-transfer validation was performed by training classifiers on Batch 1 proxy latent dimension and evaluating on Batch 2 (same subjects, independent experimental batch using the same subjects), assessing whether the learned features generalize across experimental conditions.

### 2.9 Neuronal proteins detection in brain organoids extracts spiked serum

Brain organoids (BO) were generated and protein homogenates extracted as previously described^44^. To probe (GT)_6_-SWCNT detection of NfL BO homogenates were spiked into human control serum at various concentrations (10 µg/mL, 1 µg/mL and 0.1 µg/mL). (GT)_6_-SWCNT sensor was incubated for 6 hours before protein corona extraction as previously described^45^. Briefly, after SWCNT-PC complexes underwent 3 cycles of washing and spin pull-down steps to discard unbound proteins, bound proteins were eluted from nanoparticles by heating at 95°C for 10 min in SDS/BME reducing buffer (2% SDS, 5% β-mercaptoethanol, 50 mM Tris-HCl). Protein extracts were resolved by SDS-PAGE on Mini 4 − 20% Novex Tris-Glycine Gels (Thermo Fisher). NfL (Proteintech, 12998-1-AP), NfH (Proteintech, 18934-1-AP) and Tau (Invitrogen, PA5-29610) antibodies were incubated overnight at 4 °C. Secondary antibodies conjugated with IRDye® infrared fluorophores (LI-COR) were incubated for 1 h at room temperature. Blots were visualized using the Odyssey Infrared Imaging System (LI-COR).

### 2.10 Statistical analysis

All statistical analyses were performed in Python 3.9+ using scipy, statsmodels^31^, scikit-learn^43^, and pingouin. Multiple testing correction used Benjamini–Hochberg FDR throughout unless otherwise specified. PERMANOVA^29^ and PERMDISP^30^ were implemented with custom permutation code (9,999 permutations). Linear mixed models were fit using statsmodels^31^ with maximum likelihood estimation. Effect sizes are reported as partial η² for RM-ANOVA and Cohen’s *d*_z_ for paired comparisons. Confidence intervals (95%) were computed via bootstrap (2,000 resamples). Significance threshold was set at α = 0.05 unless noted.

## 3. Results

### 3.1 Multi-chirality sensor produces partially independent temporal dynamics

Our sensing strategy relies on two established features of SWCNT–protein corona systems: the temporal evolution of the protein corona during competitive adsorption^13,14^ and chirality-dependent differences in nanotube physicochemical properties that can influence adsorption kinetics and spectral response^15–17,46–48^. We therefore first asked whether the 12 SWCNT chiralities in our sensor array exhibited time-dependent and partially non-redundant spectral dynamics after serum exposure.

We first examined global temporal structure using principal component analysis (PCA) on the 12-chirality peak-intensity vectors across all samples and timepoints. The PCA revealed a clear temporal trajectory along PC1, indicating that protein corona formation produces a coordinated shift in the overall spectral state of the multi-chirality sensor (**Figure 3d**). Paired comparisons of PC1 scores showed significant shifts between consecutive timepoints: 0 h to 6 h (mean Δ = −1.15, Cohen’s dz = −0.61, Holm-corrected p < 0.001), 6 h to 24 h (Δ = −1.87, dz = −0.86, p < 10^-5^), and 0 h to 24 h (Δ = −3.02, dz = −1.90, p < 10^-14^). These results indicate that serum exposure drives a substantial time-dependent remodeling of the sensor’s spectral state.

**Figure 1.**
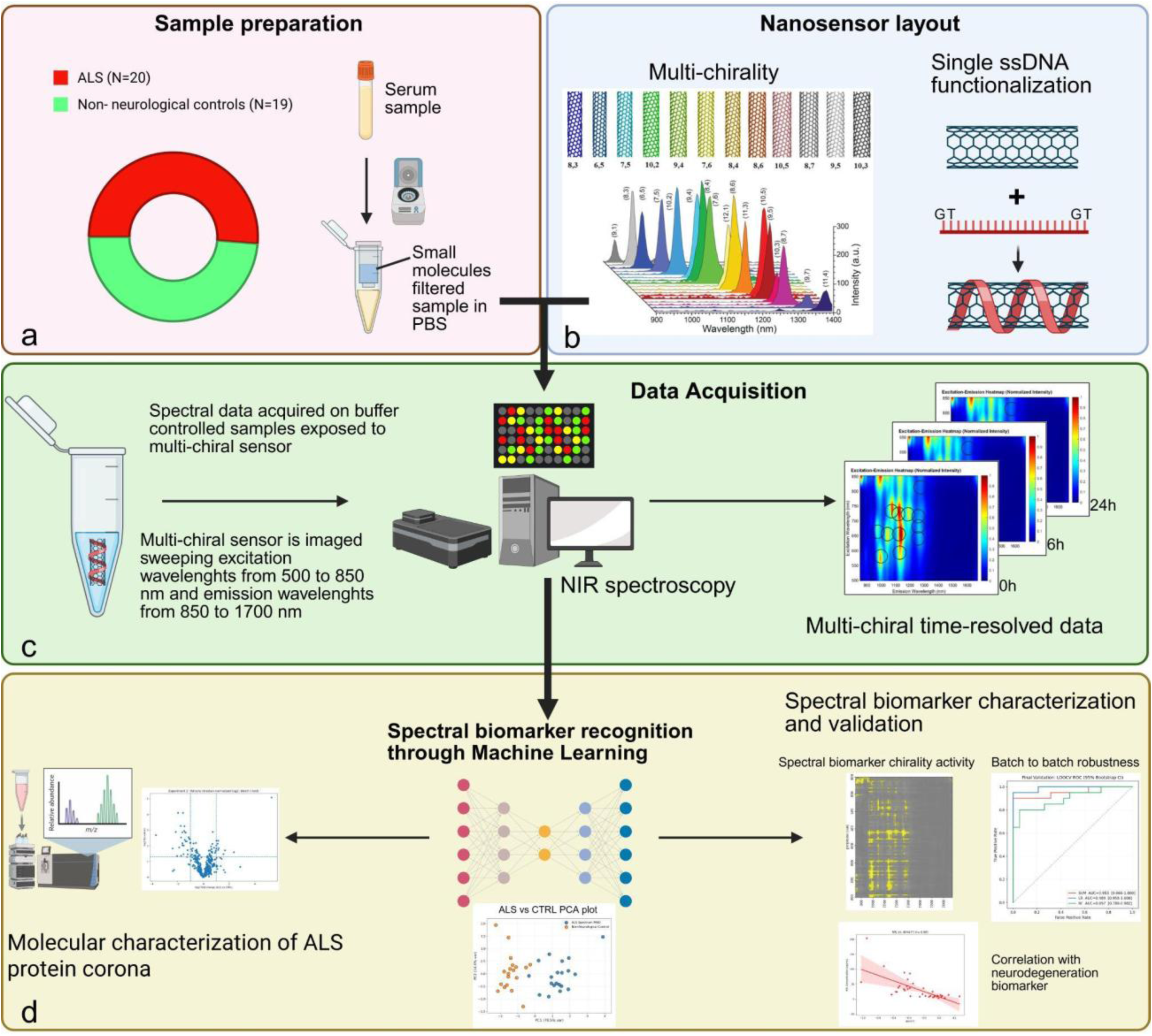
Schematic illustration of the paper outline. **a**) Before sample exposure to the SWCNT sensor, samples are pre-processed through spin column filtration to remove small molecules and homogenize protein containing buffer to improve technical reproducibility. **b**) A mixture of 12 different chiralities (6,5 ; 7,5 ; 7,6 ; 8,3 ; 8,4 ; 8,6 ; 8,7 ; 9,4; 9,5 ; 10,2 ; 10,3 ; 10,5) is functionalized with (GT)_6_ ssDNA to create the SWCNT sensor. **c**) the SWCNT sensor is incubated in the serum filtered samples and imaged using NS NanoSpectralyzer which is able to acquire a continuous emission spectrum from 850 to 1700 nm sweeping laser excitation wavelengths in steps of 5 nm from 500 to 850 nm. The image acquisition at 3 different time points (0, 6, 24 hours) generates multi-chirality time-resolved data for each sample. **d**) To decode these highly complex data is necessary a sophisticated ML technique called convolutional autoencoder. The convolutional autoencoder is able to distill a few complex spectral features (the spectral biomarker) descriptive of the ALS state. The spectral biomarker is then validated through batch-to-batch reproducibility assessment and characterized looking at its spectral and molecular determinants through importance attribution and mass spectrometry.

**Figure 2.**
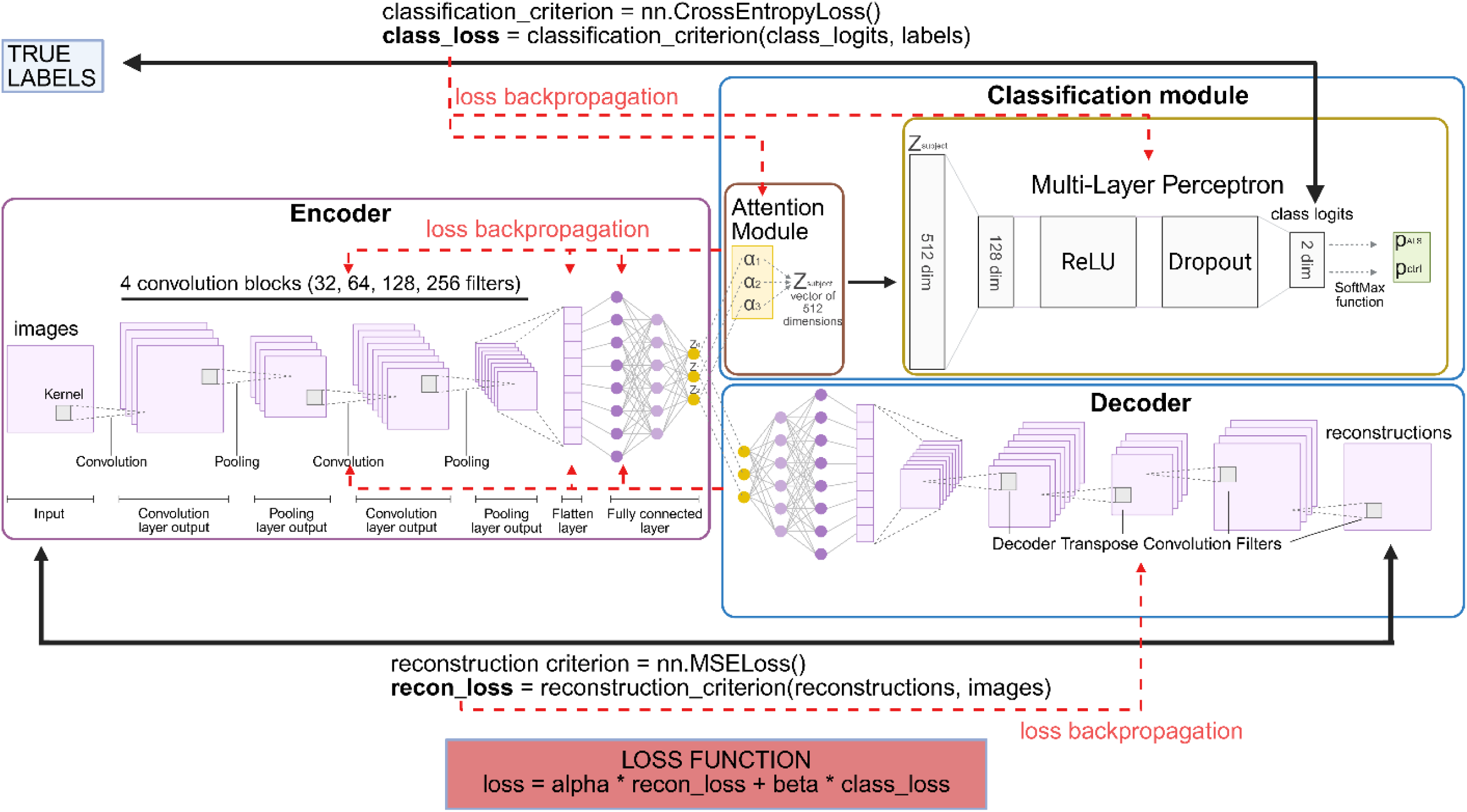
Schematic illustration of the dual-objective convolutional autoencoder architecture. The network processes time-resolved SWCNT excitation-emission matrices (EEMs) at 0h, 6h, and 24h. *Forward pass:* The encoder processes each timepoint independently through four sequential convolutional blocks with channel progression 32 → 64 → 128 → 256. The first three blocks include max pooling, and the final feature map is flattened and passed through a fully connected bottleneck layer to yield three 512-dimensional latent vectors (z_0h, z_6h, z_24h). The Decoder branch mirrors the encoder using transpose convolutions to reconstruct the original EEMs from their respective individual latent vectors. Simultaneously, the Attention & Classification Module uses a learnable attention mechanism (with Softmax normalization) to calculate importance weights (α_i) and combine the timepoints into a single aggregated subject vector (z_subject). This vector is passed to the Classification Head, implemented as a Multi-Layer Perceptron (MLP) containing a 128-node hidden layer (with ReLU and Dropout) and a final output layer producing raw logits for ALS vs. Control diagnosis. *Training and Backpropagation*: The model is optimized via a composite Loss Function summing the Mean Squared Error (MSE) of reconstruction (L_reconstruction) and the Cross-Entropy loss of classification (L_classification), weighted by αand β (both set to 1.0). As indicated by the dashed red lines, error signals backpropagate from both objectives. The classification error tunes the MLP classifier, attention weights, and the shared encoder. The reconstruction error tunes the decoder and the shared encoder. This joint optimization forces the shared convolutional encoder to extract latent features that are simultaneously discriminative for disease classification and robust enough to reconstruct the physical input data.

**Figure 3.**
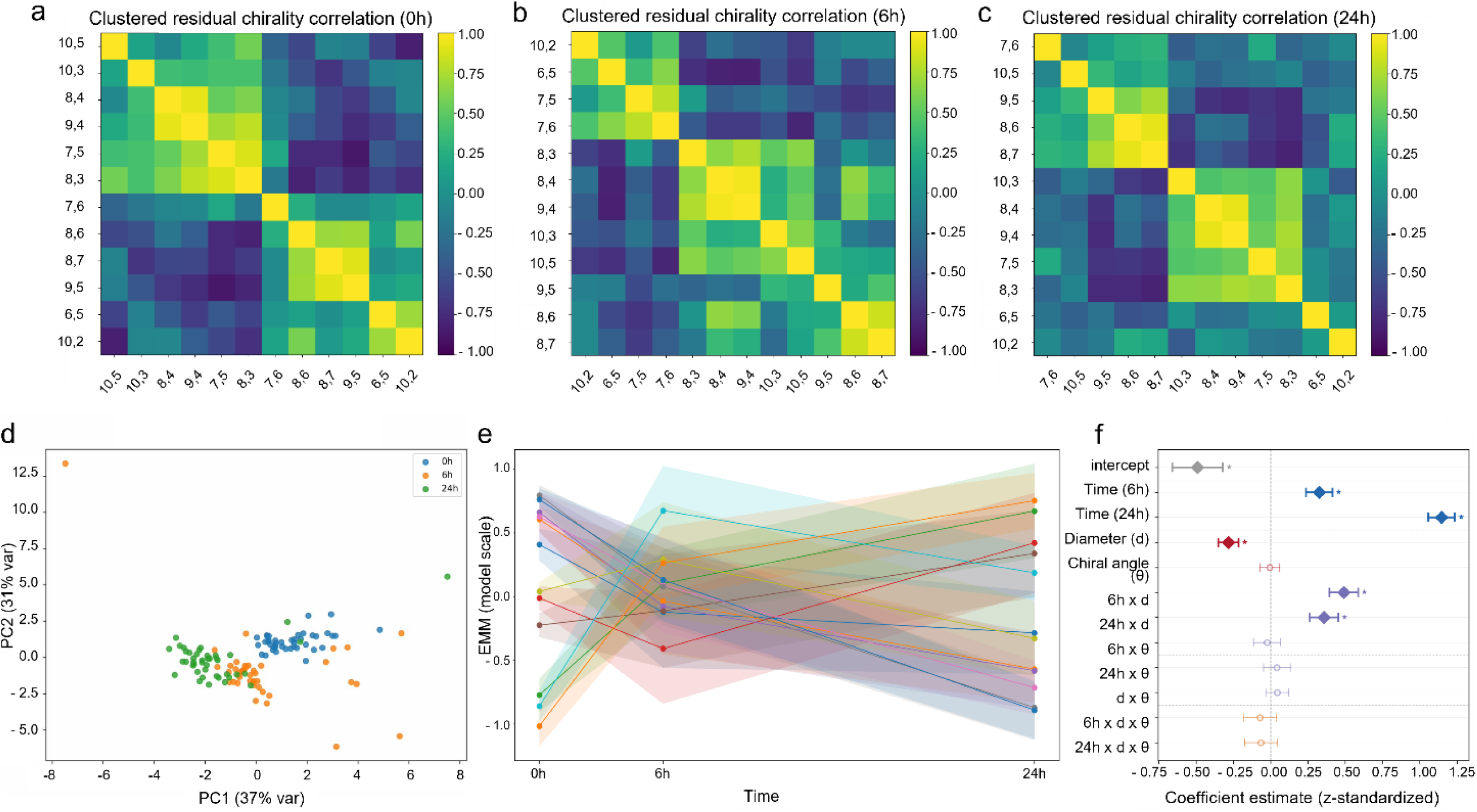
Multi-chirality temporal dynamics. ***(a–c)*** Inter-chirality Pearson correlation clustered matrices at 0 h, 6 h, and 24 h, after common mode removal, showing progressive decorrelation as protein corona matures. ***(d)*** PCA scatter colored by timepoint, demonstrating separation along PC1 with significant shifts between consecutive timepoints (paired *t*-tests, all Holm-corrected *p* < 0.001). ***(e)*** Estimated marginal means (±95% CI) from the linear mixed model for each chirality across timepoints, demonstrating heterogeneous temporal trajectories and the significant time × chirality interaction. In the graph each colored line represents the temporal trajectory of a single chirality. **(f)** Coefficient forest plot LMM fixed-effect coefficients with 95% CIs. The large positive Time (24h) coefficient (β ≈ 1.15) and the strong 6h×d and 24h×d interactions (β ≈ 0.49, 0.36) are clearly significant, while chiral angle (θ) and all its interactions have CIs crossing zero confirming intensity is purely diameter-governed.

We next quantified whether individual chiralities carried redundant or partially independent information. At baseline, peak-intensity responses were highly correlated across chirality pairs (mean |r| = 0.94; 98.5% of pairs with |r| > 0.8), consistent with a strong shared response immediately after serum application. However, inter-chirality correlations decreased as the protein corona matured: mean |r| declined to 0.87 at 6 h (81.8% of pairs with |r| > 0.8) and to 0.79 at 24 h (63.6% of pairs with |r| > 0.8) (**Table 1** and **Supp. Figure 2**). Thus, although the sensor retained a common temporal component, chirality responses became progressively less redundant over time.

Because part of this covariance could reflect a shared intensity drift affecting all chiralities, we removed the per-observation mean across all chiralities (i.e. common-mode removal) and recalculated the correlations. This markedly reduced inter-chirality correlation at all timepoints: mean |*r*| decreased from 0.94 to 0.44 at 0 h, from 0.87 to 0.45 at 6 h, and from 0.79 to 0.39 at 24 h (Table 1). Likewise, the fraction of highly correlated pairs (|*r*| > 0.8) dropped from 98.5% to 10.6% at 0 h and to below 7% at the later timepoints. When temporal dynamics were summarized as linear trend and curvature contrast scores, common-mode-removed correlations remained modest (mean |*r*| = 0.28 for linear trends and 0.39 for curvature dynamics). Pairwise significance testing (FDR-corrected) identified that 22 of 66 chirality pairs for linear trends (20 aft er common-mode removal) and 48 of 66 pairs for curvature dynamics (42 after common-mode removal) showed statistically significant correlations (FDR < 0.05). Together, these analyses indicate that the sensor contains both shared and chirality-specific temporal structure, with substantial non-redundant information.

We then tested whether the three incubation timepoints represented distinct multivariate sensor states. Global PERMANOVA^29^ on the 12-chirality feature vectors confirmed that spectral profiles changed significantly over time (F = 23.05, R² = 0.27, p < 0.0001). Pairwise comparisons showed a progressive increase in temporal separation: the 0 h vs. 6 h shift was significant but modest (F = 2.98, R² = 0.035, p = 0.013), whereas the 6 h vs. 24 h (F = 9.41, R² = 0.10, p < 0.001) and 0 h vs. 24 h contrasts (F = 33.13, R² = 0.29, p < 0.001) were substantially larger after Holm correction. This pattern persisted after common-mode removal (global F = 17.46, R² = 0.22, p < 0.0001), confirming that temporal remodeling was not solely driven by shared intensity drift (**Supp. Figure 3a**). PERMDISP analysis^30^ further showed that multivariate dispersion changed over time (Friedman χ² = 11.76, p = 0.003). Dispersion was lowest at 0 h, increased significantly by 6 h (Wilcoxon p < 0.001), and partially contracted by 24 h while remaining elevated relative to baseline (0 h vs. 24 h, p = 0.005; 6 h vs. 24 h, not significant; **Supp. Figure 3b**). This pattern suggests a two-phase corona response: an initial expansion in inter-sample spectral variability followed by partial stabilization at 24 h.

Finally, we wanted to *assess chirality specific temporal dynamics*. Per-chirality temporal changes were quantified using repeated-measures ANOVA (RM-ANOVA) with Benjamini–Hochberg FDR correction across the 12 chiralities, followed by paired post-hoc comparisons^25^. Per-chirality RM-ANOVA demonstrated that all the 12 chiralities exhibited significant temporal changes (all FDR-corrected *p* < 0.05, **Supp. Figure 3c**). The largest effects were observed for chirality (9,5) (*F* = 62.14, partial η² = 0.60), (10,5) (*F* = 52.97, η² = 0.56), and (8,7) (*F* = 39.98, η² = 0.49), while the smallest was (9,4) (*F* = 4.84, η² = 0.11). To further investigate the multiplexing effect of chirality and time on the spectral response, we used a linear mixed model (LMM, **Figure 3e**). Critically, the LMM analysis revealed a highly significant time × chirality interaction (*p* < 0.0001), further proving that different chiralities evolve at different rates, a fundamental assumption of our multiplexing strategy. Post-hoc contrasts (FDR-corrected) showed that 28 of 36 chirality–timepoint comparisons were significant (FDR < 0.05), with the 24 h–0 h contrast significant for all 12 chirality (**Supp. Figure 3d**). Together, these results demonstrate that our multi-chirality sensor generates a rich, time-evolving spectral landscape with partially independent dynamics across chirality, providing the information basis for discriminative feature extraction.

Finally, we asked which structural properties of the nanotubes explained these chirality-specific dynamics. We fitted linear mixed models parameterizing each chirality by tube diameter and chiral angle for peak fluorescence intensity (**Figure 3f**) and emission center wavelength (**Supp. Figure 4b**). For peak fluorescence intensity, likelihood ratio tests^32^ revealed that time and diameter were the dominant determinants, while chiral angle played a negligible role. Time explained the largest fraction of variance (LR = 629.9, df = 8, p < 10^-15^), with fluorescence increasing progressively from 0 h to 6 h (β = 0.33, p < 0.001) and more substantially to 24 h (β = 1.15, p < 0.001). Diameter was the sole significant geometric predictor (LR = 111.1, df = 6, p < 10^-15^), showing a negative main effect (β = −0.28, p < 0.001), indicating that larger-diameter tubes display lower baseline fluorescence intensity. Critically, the time × diameter interactions were large and positive (β_6h = 0.49, p < 0.001; β_24h = 0.36, p < 0.001), meaning that larger-diameter tubes undergo a stronger fluorescence increase during corona formation, effectively converging toward the intensity levels of smaller tubes. In contrast, chiral angle contributed negligibly to fluorescence intensity variation (LR = 4.3, df = 6, p = 0.64), and the three-way interaction was also non-significant (LR = 2.0, df = 2, p = 0.37). The model’s intraclass correlation coefficient (ICC = 0.35)^49^ indicated that approximately one-third of the total variance in fluorescence intensity is attributable to differences between different subjects, confirming meaningful sample-level signal in the intensity channel. The emission center wavelength model revealed a fundamentally different and richer pattern: both diameter and chiral angle were highly significant, and the full three-way interaction emerged as statistically meaningful. Time again explained the largest variance component (LR = 173.4, df = 8, p < 10^-15^), but with an asymmetric temporal profile: the 0 h to 6 h shift was substantial (β = 0.40, p < 0.001), while the 0 h to 24 h shift was smaller and only marginally significant (β = 0.11, p = 0.073), indicating that most solvatochromic shifting occurs during the early phase of corona formation^48^ (**Supp. Figure 4b**). Both structural parameters contributed significantly: diameter (LR = 74.6, df = 6, p = 4.7 × 10^-14^; β = −0.28, p < 0.001) and chiral angle (LR = 68.1, df = 6, p = 1.0 × 10^-12^; β = −0.23, p < 0.001), each showing negative main effects—meaning that tubes with larger diameter or greater chiral angle exhibit blue-shifted emission centers. Both structural parameters also showed strong positive interactions with time: for diameter, β_6h = 0.38 and β_24h = 0.48 (both p < 0.001); for chiral angle, β_6h = 0.40 and β_24h = 0.29 (both p < 0.001). Notably, the time × diameter × chiral angle interaction was significant (LR = 19.3, df = 2, p = 6.6 × 10^-5^), driven by a strong negative coefficient at 24 h (β = −0.26, p = 4.5 × 10^-4^). This interaction suggests that late-stage solvatochromic shifts are constrained by nanotube geometry, although the physical basis of this constraint will require further validation. The lower ICC for emission center (0.07 vs. 0.35 for intensity) further confirms that emission wavelength is primarily determined by nanotube electronic structure and the local dielectric environment rather than sample-level variability, consistent with its role as a reporter of corona composition rather than total protein loading.

### 3.2 Convolutional autoencoder extracts discriminative latent features converging on EEM regions of SWCNT spectral activity

To fully exploit the complex multi-chirality temporal information, we trained our designed five-fold convolutional autoencoder model. The model achieved a mean fold accuracy of 84.6 ± 5.4% and a mean fold AUC of 0.85 ± 0.14 across the five subject-level validation folds. The pooled out-of-fold performance was consistent with these fold-level estimates, with an overall accuracy of 84.6% and AUC = 0.87 (**Supp. Figure 5**).

The learned attention mechanism assigned the highest mean weight to the 6 h timepoint, followed by 24 h and 0 h (α_6_ₕ = 0.389 ± 0.28, α₂₄ₕ = 0.331 ± 0.28, α₀ₕ = 0.281 ± 0.25; mean ± SD across held-out subjects; **Supp. Figure 6**). At the class level, control samples showed higher mean weighting of the 6 h state (CTRL: 0.450 ± 0.28; ALS: 0.330 ± 0.26), whereas ALS samples showed a more evenly distributed attention profile across the three timepoints. Because the between-class differences in attention weights were modest relative to inter-subject variability, we interpreted the attention analysis primarily as evidence that intermediate and late corona states contributed substantially to classification, rather than as an independent class-separating feature. This pattern is consistent with the interpretation that serum composition alters the trajectory of corona maturation rather than producing a purely static spectral difference.

We then asked whether the model relied on spectroscopically meaningful regions of the EEMs rather than arbitrary image artifacts. First, we examined reconstruction error across folds and timepoints. Average per-pixel MSE maps showed structured reconstruction behavior, with low-error regions overlapping multiple known chirality-associated excitation–emission coordinates (**Figure 4a–c**). This indicates that the autoencoder learned to preserve major SWCNT spectral features during reconstruction.

**Figure 4.**
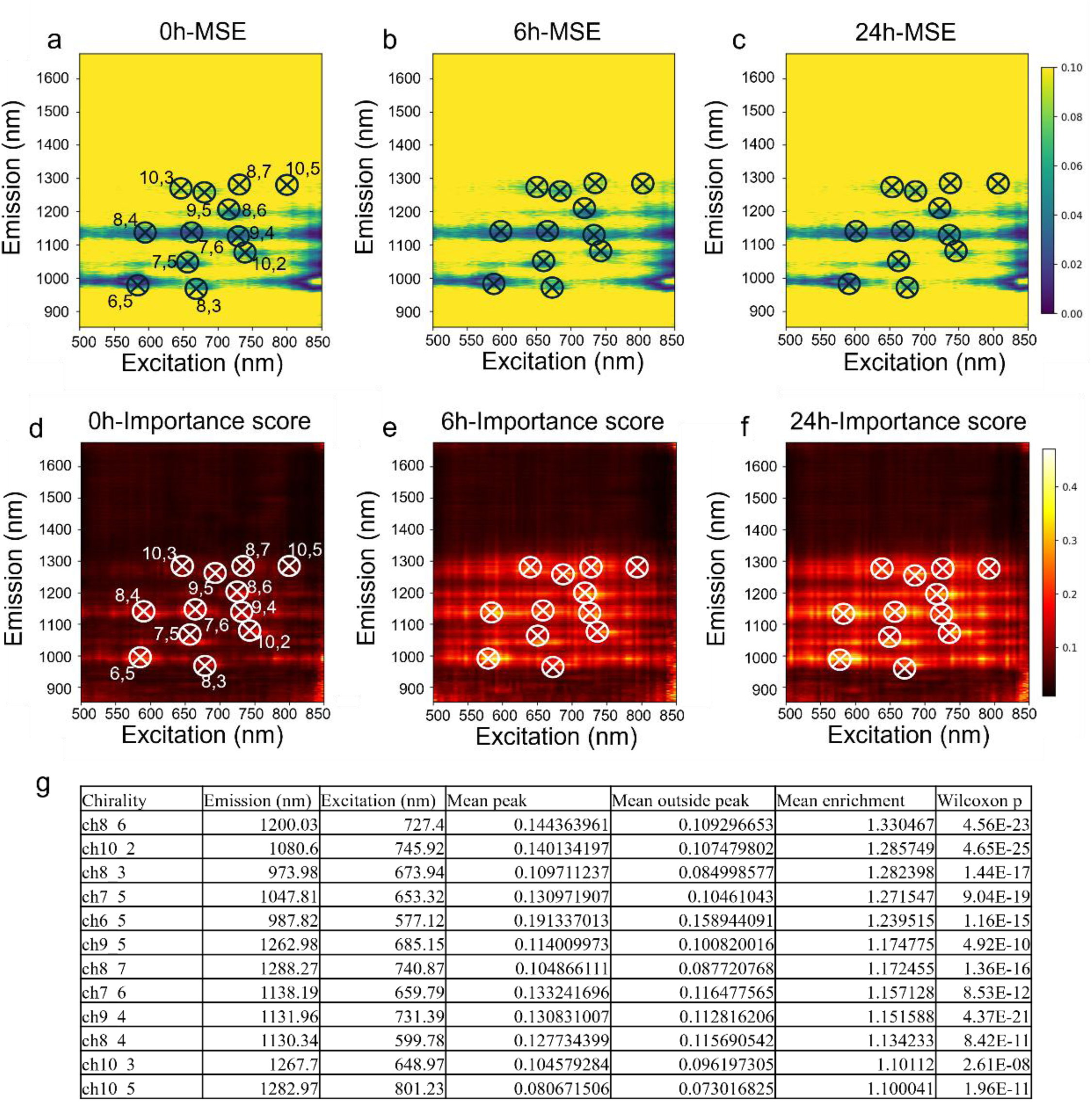
Autoencoder interpretability analysis. ***(a-c)*** Per time-point reconstruction error map (average of 5 models) showing per-pixel MSE distribution. ***(d-f)*** Per time-point importance score map (average of 5 models) integrating gradient saliency and linear probing for the latent dimension, projected onto the excitation–emission coordinate space. Black crosses in a-c indicate the precise excitation-emission coordinate of each chirality peak. ***(g)*** Summary table of chirality peak importance score. For each chirality the importance score computed for each latent dimension is significantly higher in the proximity of the chirality peak (Mean peak column) than in the neighboring pixels (mean outside peak column).

Second, we examined model attribution using gradient saliency^40^, integrated gradients^41^, and linear probing^42^. Consensus attribution maps converged on spectral regions spanning multiple chirality-associated excitation–emission coordinates (**Figure 4d–f**), indicating that the latent representations were informed by distributed SWCNT spectral activity rather than by isolated background regions. To quantify this enrichment, we compared the importance score around each chirality peak with the importance score in its surrounding neighborhood. For each of the top 10 univariate AUC-ranked latent dimensions from each of the five models across the three timepoints, we calculated mean importance within a local chirality-peak ROI and compared it with the surrounding ring region. Across all 12 chiralities, the average peak-region importance exceeded the neighboring background region, yielding enrichment ratios greater than 1 for every chirality (**Figure 4g**). Together, these results show that the convolutional autoencoder learned representations that were both discriminative for ALS status and anchored in physically meaningful SWCNT spectral regions. The convergence of reconstruction-error and attribution analyses on known chirality-associated coordinates supports the interpretation that the model extracted biologically relevant spectral structure from the multi-chirality sensor, rather than relying on nonspecific image artifacts.

### 3.3 Autoencoder-generated features achieve high classification performance and resist batch effects

Having shown that the convolutional autoencoder learned spectroscopically meaningful representations, we next asked whether its 512-dimensional latent space could be reduced to a compact set of interpretable spectral biomarker candidates. To do this, we applied the two-stage latent-dimension selection and frozen-weight proxy reconstruction pipeline described in Methods 2.8 (**Supp. Figure 1**). This workflow first identifies discriminative, non-redundant latent dimensions from the five cross-validated autoencoder models and then reconstructs their values analytically using frozen encoder and attention weights, without fitting additional parameters during proxy extraction. From the full candidate pool of 2,560 latent features (512 dimensions × 5 folds), AUC-based ranking, stability filtering, Spearman redundancy pruning (|ρ| < 0.85), and iterative LOOCV classifier evaluation selected three non-redundant latent dimensions: dim477 from fold 3, dim103 from fold 1, and dim370 from fold 3. These dimensions showed high univariate ALS-versus-control discrimination, with AUC values of 0.987 for dim477, 0.979 for dim103, and 0.978 for dim370 (**Figure 5d** and **Supp. Figure 7**). Permutation testing of the final three-feature set confirmed that this discrimination was unlikely to arise by chance. The per-timepoint contribution analysis revealed that these three dimensions encode distinct temporal patterns. dim477 was most strongly correlated with the 24 h timepoint (*r*_24h_ = −0.39) and received the highest attention weight at 24 h (α_24h_ = 0.55), reflecting equilibrium corona composition. Dim103 and dim370, in contrast, drew information from both 6 h and 24 h timepoints (*r*_6h_ = 0.54 and 0.44; *r*_24h_ = 0.53 and 0.65, respectively), capturing corona exchange dynamics. The 0 h timepoint contributed minimally to all three dimensions (|*r*_0h_| < 0.07), consistent with the pre-incubation snapshot carrying limited discriminative information on its own. Strikingly dim477 by itself represents a great candidate as spectral biomarker classifying correctly almost all the samples despite having been derived from just 30 of them (model 3). Interestingly, the importance score map of dim477 revealed that the pixel importance is almost homogeneously distributed across the 12 chirality (**Figure 5c**) suggesting the latent dimension extracts classification-useful information from all the components of the sensor.

**Figure 5.**
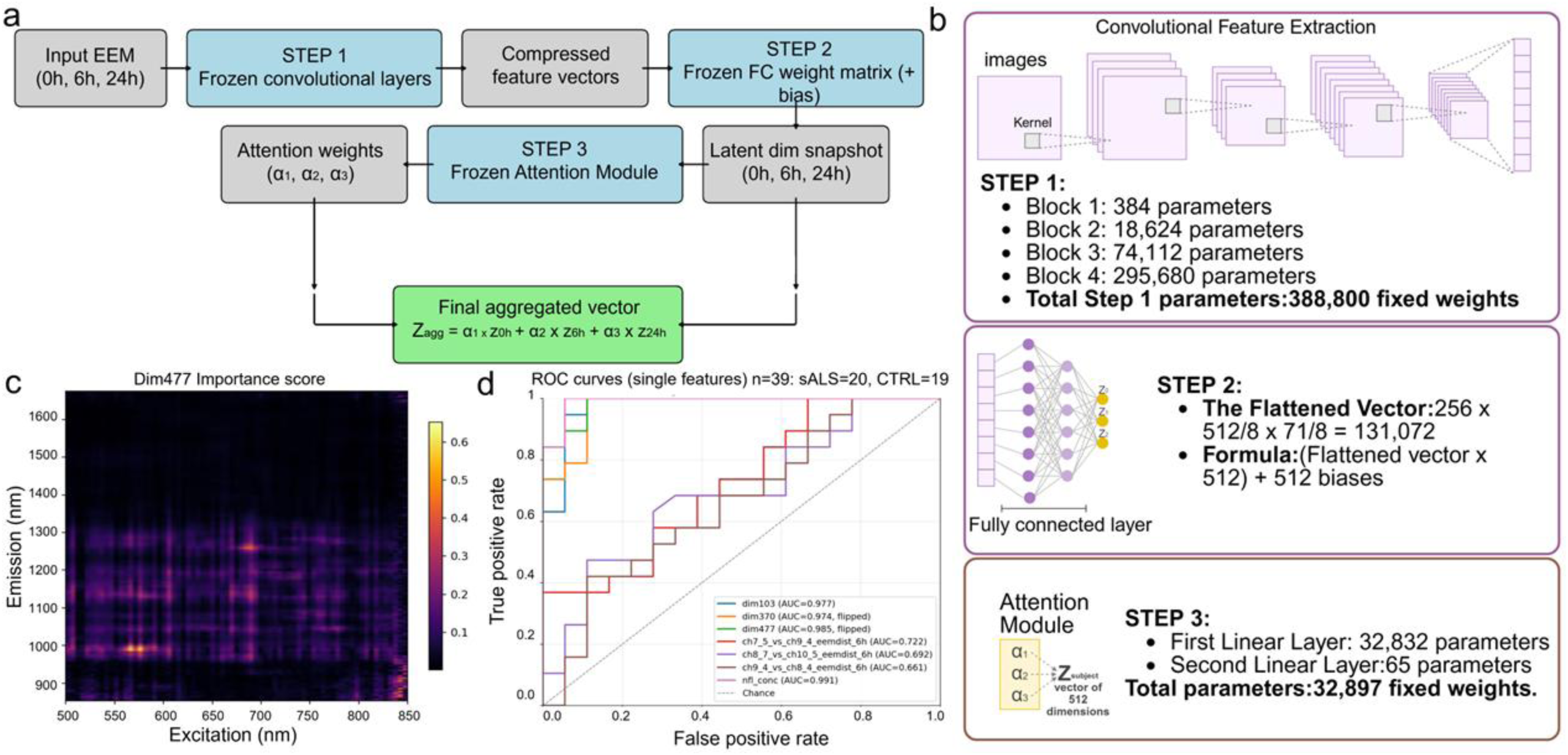
Computation of spectral biomarkers from the spectral maps. ***(a)*** Schematic representation of the latent dimension derivation workflow from the three input images using the trained autoencoder derived weights. ***(b)*** Illustration of the latent dimension computational steps referring to the convolutional autoencoder architecture illustrated in Figure 2. ***(c)*** Importance score map of dim477. **(d)** ROC curve comparison across the top 3 univariate ranked latent dimensions (dim370 in yellow, dim103 in blue and dim477 in green) the top 3 univariate ranked physical pairwise features (ch7_5_vs_ch9_4_eemdist_6h in red, ch8_7_vs_ch10_5_eemdist_6h in purple and ch9_4_vs_ch8_4_eemdist_6h in brown) and NfL concentration (pink line). For all the top 3 ranked latent dimensions the AUC (dim370 = 0.978, dim103 = 0.979 and dim477 = 0.987) is greater than the AUC of physical pairwise features (ch7_5_vs_ch9_4_eemdist_6h = 0.737, ch8_7_vs_ch10_5_eemdist_6h = 0.717 and ch9_4_vs_ch8_4_eemdist_6h = 0.689). The “flipped” suffix on dim477 and dim370 indicates that the sign of these dimensions was inverted prior to ROC computation: because the polarity of autoencoder latent dimensions is determined by random weight initialization and carries no biological directionality, the sign is conventionally chosen so that higher values correspond to the ALS (positive) class.

We then reconstructed these selected latent dimensions as deterministic proxy features using the frozen autoencoder weights. Briefly, each sample’s EEM at each timepoint was passed through the trained convolutional encoder, the resulting feature representation was projected through the frozen fully connected bottleneck layer to obtain a 512-dimensional latent vector per timepoint, and the frozen attention network produced sample-specific weights for aggregating the 0 h, 6 h, and 24 h latent snapshots. The target latent dimensions were then extracted from this attention-weighted aggregated vector. Because no new parameters were fit during this proxy extraction step, the resulting features represent fixed readouts of the trained encoder rather than newly optimized classifiers. Using the 3 proxy latent dimensions, a linear SVM achieved AUC = 0.98 in LOOCV (36/39 correct), while both RF and LR reached AUC ≥ 0.99 and 97.4% accuracy (38/39 correct; Table 2). These results indicate that the learned representation can be compressed from the full EEM image sequence into a small set of highly discriminative latent biomarker candidates.

We next compared the autoencoder-derived features with conventional hand-engineered spectral descriptors. For each chirality and timepoint, we extracted fitted peak amplitude, emission center wavelength, excitation center wavelength, FWHM, area under the curve, skewness, kurtosis, and fit-quality metrics, together with pairwise chirality-ratio and peak-distance descriptors. In total, this yielded 648 per-chirality descriptors and 396 pairwise descriptors per sample. We designed a 5 folds approach similar to the one used for the convolutional autoencoder. Inside each fold SVM, LR and RF were trained and validated using LOOCV on 31 samples. To avoid any data leakage, feature selection was done inside the LOOCV based on F-ANOVA and a correlation filter. The best-performing algorithm was RF achieving an AUC of 0.67. The pipeline strongly converged on a single dominant feature: ch7_5_vs_ch9_4_eemdist_6h (the EEM distance between chirality (7,5) and (9,4) at 6h), selected in 4/5 folds for all three classifiers. To further compare physical descriptors with latent dimensions we used F-ANOVA to rank physical descriptors in the full dataset. Strikingly 72% (32/44) of the features with p-value < 0.05 belong to the group of the pairwise features. These features represent the impact of sample’s protein composition on inter-chirality relationships (intensity ratios and eem distances) which better describe the sensor system evolution rather than describing single chirality response. Indeed, denoising the dataset removing the 648 per-chirality descriptors yielded to a higher classification accuracy (RF: 0.639±0.149) and improved the AUC (RF: 0.767±0.098) in a pairwise features only dataset. These findings further confirm the need of a system wise approach, as our convolutional autoencoder architecture, to capture complex sensor dynamics informative of the phenotypic state of the sample. To prove the superior capability of our convolutional autoencoder to capture these complex features we compared the ROC curves of the best ranked autoencoder and pairwise features. The striking difference between the AUC of dim477 (AUC=0.97) and ch7_5_vs_ch9_4_eemdist_6h (AUC=0.73) highlights the superiority of the autoencoder derived non-linear complex features over fitting-based computed features (**Figure 5d** and **Supp. Figure 7**). Notably, the best latent dimension (dim477) reached the AUC level of Neurofilament Light chain (NfL), a well-known neurodegenerative biomarker measured with SIMOA (Quanterix)^20^, a high sensitivity immunoassay.

We assessed whether the selected proxy latent dimensions were robust to experimental batch effects. Classifiers were trained on Batch 1 proxy latent dimensions and evaluated on Batch 2 (same subjects, independent experimental session). The SVM achieved 97.4% accuracy (38/39 correct) on Batch 2, with RF and LR both achieving 94.9% (37/39). Since Batch 2 used the same biological subjects, this experiment does not constitute independent cohort validation; rather, it tests technical and experimental reproducibility of the learned spectral features. The high transfer performance indicates that the selected latent dimensions are stable across independent acquisition batches and are unlikely to reflect batch-specific artifacts.

Finally, we performed extensive sensitivity analyses to exclude confounding (**Supp. Table 2**). Age, the only demographic imbalance (ALS 58 vs. control 49 years, p = 0.026), did not drive the signature: it was uncorrelated with the latent features within each group, discrimination was retained in a 1:1 age-matched subset (p = 0.91; AUC = 0.99), and retraining the model on that age-balanced cohort reproduced the separation with no loss beyond the expected sample-size effect. Sex, ethnicity, and race were balanced between groups, and within the ALS arm the signature was independent of disease-modifying therapy (riluzole, edaravone), disease severity, duration, and genetic subtype, indicating a disease-state marker rather than a treatment or severity readout. Gross fluorescence intensity did not discriminate the groups (AUC = 0.52), arguing against a batch or brightness artifact. Two controls carried a C9orf72 expansion; their exclusion left classifier performance unchanged and, across latent representations, neither was significantly more ALS-like than other controls (p ≥ 0.6)—though the dim477 feature did classify the older carrier (age 59, but not the 37-year-old) as the most ALS-like control, a single-representation observation compatible with a presymptomatic signal that warrants follow-up.

Taken together these findings highlight the capacity of our convolutional autoencoder’s architecture to discover complex relationships between spectral data that can be translated into a candidate spectral biomarker as we illustrated with dim477.

### 3.4 SWCNT spectral features covary with serum NfL concentration

To investigate whether the autoencoder-derived spectral biomarker (dim477) captures information related to established ALS biomarkers, we correlated latent dimensions with serum neurofilament light chain (NfL) concentration measured by Simoa. NfL is a cytoskeletal protein released upon neuronal damage and is considered the most promising fluid biomarker for ALS, though it lacks disease specificity^4,50,51^. Dim477 showed a significant inverse association with serum NfL concentration across the full cohort (r = −0.68, p < 0.001; **Figure 6a**), indicating that the learned SWCNT spectral representation covaries with a recognized marker of neuronal injury.

**Figure 6.**
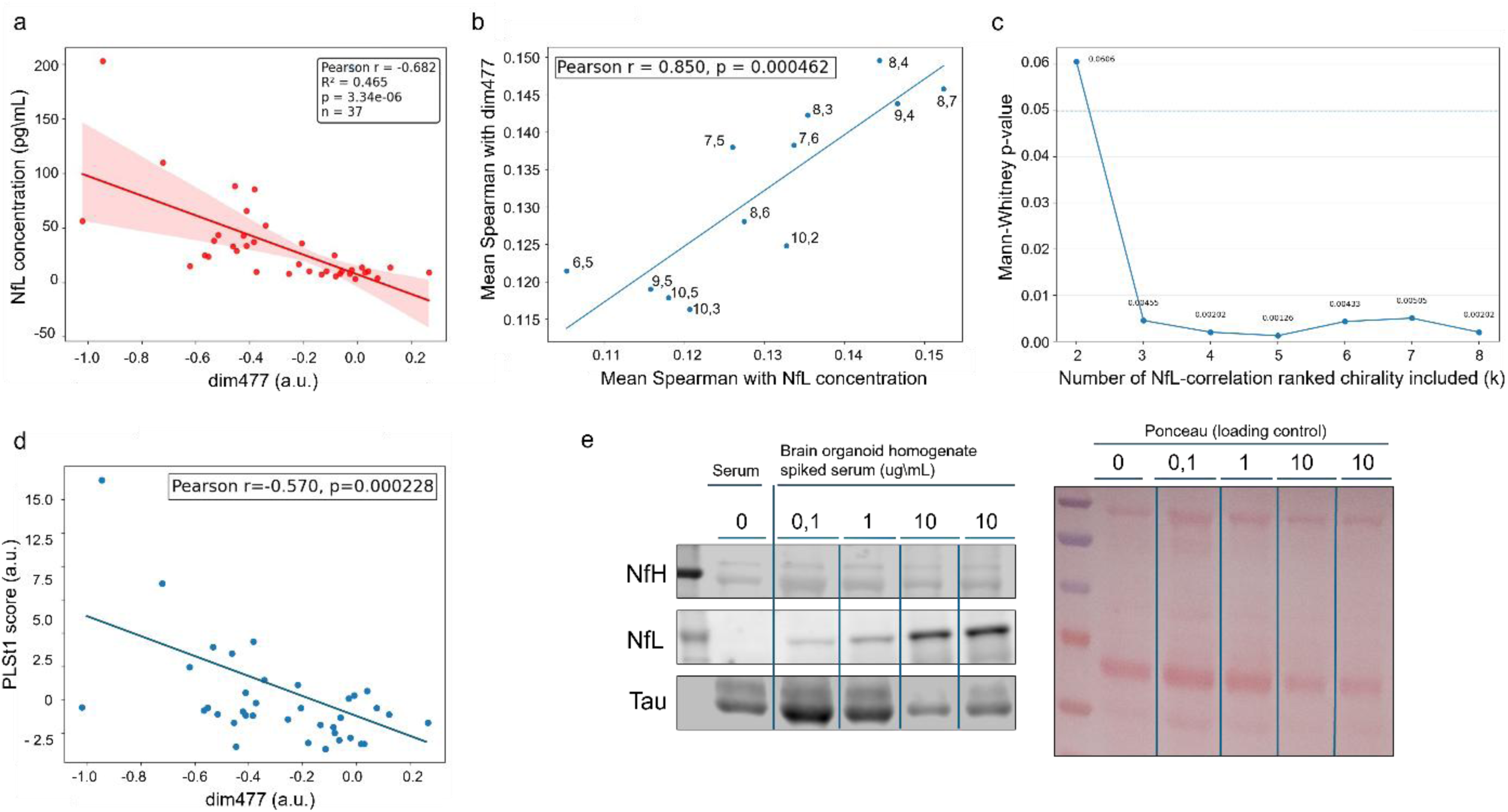
Neurofilament light chain correlation with autoencoder features. ***(a)*** Scatter plot showing the correlation between proxy latent dim477 and serum NfL concentration (r = −0.68, *p* < 0.001). ***(b)*** Plot of mean Spearman of grouped chirality features with dim477 on y-axis and mean Spearman of grouped chirality features with NfL on x-axis. The plot shows how the increment of mean Spearman of grouped chirality features with NfL corresponds to an increment of mean Spearman of grouped chirality features with dim477. ***(c)*** Rank-based enrichment analysis. We ranked chirality according to their mean NfL association and asked whether the higher-ranked chirality also tended to have stronger dim477 associations than the remaining chirality. This comparison was evaluated with a one-sided Mann–Whitney U test for progressive increment of rank threshold k. ***(d)*** dim477 correlation with first PLS latent score. PLS was performed using only non-pairwise chirality descriptors to model serum NfL. ***(e)*** Representative Western blot on protein corona isolated proteins of SWCNT incubated in filtered serum protein extract or spiked with different concentrations of brain organoid homogenate.

To determine whether this dim477–NfL relationship could be linked to interpretable spectral features of the nanotube response, we next examined chirality-derived descriptors extracted from the emission profiles. To determine whether the dim477–NfL relationship was reflected in the chirality structure of the spectral descriptors, we aggregated descriptor associations at the chirality level and examined whether chirality with stronger mean NfL-linked profiles also showed stronger mean dim477 associations. To this extent pairwise features were attributed to the chirality involved in the feature computation (**Figure 6b**). A significant positive relationship (r =0.85, p = 0.0004) was observed across chirality between chirality-NfL correlation mean and chirality-dim477 correlation mean, supporting the idea that Dim477 captures distributed NfL-related spectral information. Complementary, to assess whether chirality-level descriptor profiles more strongly associated with NfL also tended to show stronger association with Dim477, we performed a rank-based enrichment analysis. Descriptor correlations were first aggregated by chirality by computing, for each chirality, the mean absolute correlation of all descriptors involving that chirality with NfL and, separately, with dim477. Chirality was then ranked according to their mean NfL association. For a given rank threshold k, the mean absolute dim477 associations of the top-ranked chirality were compared with those of the remaining chirality using a one-sided Mann–Whitney U test. To reduce dependence on an arbitrary cutoff, this procedure was repeated across multiple values of k, showing that the enrichment remained significant across a broad range of thresholds (**Figure 6c**). This analysis therefore tests whether stronger NfL-linked chirality profiles are systematically accompanied by stronger dim477 associations at the chirality level. This rank-based enrichment analysis gave consistent results across multiple cutoffs, indicating that this pattern was not dependent on a single threshold choice.

We also noted that the physical features correlating the strongest with both NfL and dim477 are pairwise features (ex. ch10_2_vs_ch8_4_ratio_6h with ρ(dim477) = 0.413 and ρ(NfL) = -0.412, ch8_4_vs_ch7_6_ratio_6h with ρ(dim477) = -0.420 and ρ(NfL) = 0.340, ch8_4_vs_ch8_7_eemdist_24h with ρ(dim477) = -0.327 and ρ(NfL) = 0.366, ch8_7_vs_ch10_5_eemdist_24h with ρ(dim477) = 0.346 and ρ(NfL) = -0.332). These analyses indicated that the dim477 signal related to NfL concentration is not dominated by a single chirality-specific descriptor, but instead is distributed across multiple relational features, with the strongest signal arising from inter-chirality descriptors.

Because many descriptors are pairwise by construction and encoded relationships between two chirality, attribution to a single chirality is not possible. We therefore adopted a second complementary strategy. Indeed, to test whether interpretable single-chirality descriptors also contained NfL-related structure aligned with dim477, we applied partial least squares (PLS) regression using only non-pairwise chirality descriptors to model serum NfL. In this supervised framework, dim477 was significantly correlated with the first PLS latent score (r = -0.570, p = 2.28 × 10^-4^), showing that dim477 aligns with the dominant NfL-related axis present in the single-chirality descriptor space (**Figure 6d**). However, the NfL values predicted from this non-pairwise descriptor block were not themselves significantly correlated with dim477 (r = 0.088, p = 0.605), indicating that single-chirality descriptors alone do not fully explain the dim477–NfL relationship. These results indicate that the autoencoder captures and concentrates the NfL-related spectral information that is distributed across multiple chirality–descriptor combinations into a single latent dimension.

In line with this observation, we wanted to look at the capability of our SWCNT sensor to adsorb NfL and other neurodegeneration related (Tau protein and Neurofilament Heavy chain, NfH). To this aim we incubated (GT)_6_-SWCNT sensor with serum protein extracts enriched with brain organoid proteins at different dilutions (10 µg/mL, 1 µg/mL and 0.1 µg/mL) for 6 hours to create a brain proteome enriched protein corona. We detected by Western Blot the presence of all the three investigated proteins (NfL, Tau and NfH) at each of the different concentrations. All of them were detected at the lowest brain organoid homogenate concentration while Tau and NfH were both detectable in un-spiked native serum extracted proteins (**Figure 6e**). Together, these results indicate that dim477 covaries with serum NfL and with NfL-associated spectral structure distributed across multiple chirality relationships. However, dim477 should be interpreted as an NfL-associated multivariate corona signature rather than a direct optical surrogate for NfL concentration alone.

### 3.5 Mass spectrometry reveals ALS-associated protein corona composition

To provide a molecular basis for the spectral differences detected by the SWCNT sensor, we performed LC–MS/MS analysis of the protein corona formed on SWCNTs after incubation with ALS and Control serum protein extracts. Per group pooled protein corona samples were analyzed from three independent experiments. A total of 600 proteins were identified and quantified. Following removal of likely contaminant, hemolysis-, platelet-, and coagulation-associated proteins, the filtered serum proteomic dataset comprised 540 detected protein groups, of which 126 were differentially abundant at FDR < 0.10 (**Figure 7b**). PCA of the normalized, imputed abundance matrix revealed clear separation of ALS and control samples along PC1, which accounted for 53.8% of the total variance, whereas PC2 explained 18.6%. Group separation was evident along PC1 (Welch’s p = 0.0118) but not PC2 (p = 0.690), and between-group distances in PCA space exceeded within-group distances (p = 2.00 × 10^-4^), indicating that disease status is the major source of structured proteomic variation (**Figure 7a**).

**Figure 7.**
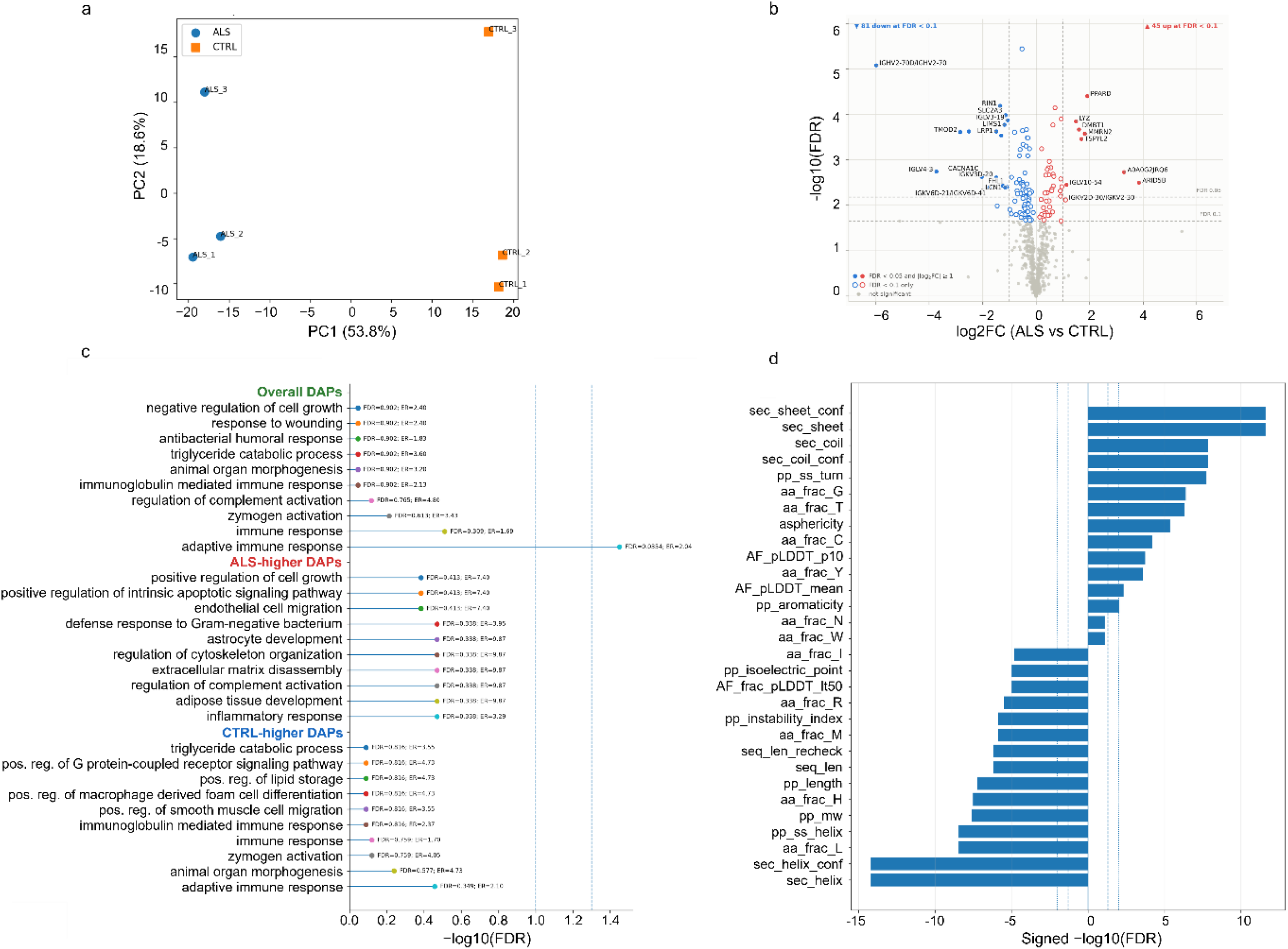
Mass spectrometry analysis of the SWCNT protein corona. ***(a)*** PCA of median-normalized log_2_ protein abundances showing separation between ALS and control corona compositions. ***(b)*** Volcano plot of differentially abundant proteins. The x-axis shows log_2_ fold change (ALS/CTRL) and the y-axis shows –log_10_ FDR-corrected *p*-value. Proteins passing FDR < 0.10 are colored orange. ***(c)*** Top 10 GO Biological Process enrichments in total DAPs, ALS up-regulated DAPs and Control up-regulated DAPs. ***(d)*** Top 30 strongest directional chemical-physical features differences between serum proteins enriched in (GT)_6_-SWCNT protein corona and non-(GT)_6_-SWCNT enriched proteins.

Gene Ontology biological process over-representation analysis was then performed using the full detected proteome as background. Among all differentially abundant proteins, adaptive immune response was the only term that remained significant after multiple-testing correction (p = 1.03 × 10^-4^, BH FDR = 0.0354), with 23 of 108 annotated differentially abundant proteins assigned to this category compared with 54 of 518 annotated proteins in the background, corresponding to an enrichment ratio of 2.04 (**Figure 7c**). No additional GO term remained significant after correction. When proteins were separated by direction of change, no GO term passed FDR correction in either subset. However, the ALS-higher proteins showed a coherent nominal enrichment profile centered on inflammatory and innate immune biology, including inflammatory response (p = 0.00645, FDR = 0.338), regulation of complement activation (p = 0.0128, FDR = 0.338), extracellular matrix disassembly (p = 0.0128, FDR = 0.338), and defense response to Gram-negative bacterium (p = 0.0140, FDR = 0.338). These terms were driven by proteins such as CFH, C3, LYZ, DMBT1, LTF, CTSG, SERPINA3, and S100A8. In contrast, the CTRL-higher proteins were nominally enriched for adaptive immune response (p = 0.00133, FDR = 0.349) and related humoral immune categories, consistent with the predominance of immunoglobulin-associated proteins in that subset. Collectively, these results indicate that the filtered ALS serum proteome remains globally distinct from controls and is characterized by a statistically robust enrichment of adaptive immune biology, together with a reproducible but non-significant trend toward innate inflammatory and complement-related processes among proteins elevated in ALS. These proteomic findings provide a direct molecular basis for the SWCNT sensor’s capacity to distinguish ALS from control serum.

We next examined whether the (GT)_6_-SWCNT corona exhibited chemical and biophysical selectivity relative to the broader serum proteome, and whether those same properties could explain differences between ALS and control coronas. To do this, proteins identified in the (GT)_6_-SWCNT corona were first compared with serum proteins present in the serum background but not detected in (GT)_6_-SWCNT. For each protein, we assembled a feature matrix combining sequence-derived descriptors calculated from the primary amino acid sequence using BioPython ProtParam/ProteinAnalysis^26^ with structure-related descriptors from AlphaFold^27,28^ and UniProt^24^ databases. These features included protein length, molecular weight, GRAVY hydropathy, aromaticity, instability index, isoelectric point, estimated charge at pH 7.4, mean flexibility, predicted secondary-structure propensities, the fractional abundance of each amino acid residue, and AlphaFold-derived confidence and shape metrics. Each feature was then tested independently between (GT)_6_-SWCNT and serum-background proteins using a two-sided Mann–Whitney U test with Benjamini–Hochberg false discovery rate correction, and effect direction was summarized by the rank-biserial correlation (RBC). This analysis showed that the (GT)_6_-SWCNT corona was not a random subset of serum proteins but instead displayed a distinct chemical signature. The strongest differences were structural: (GT)_6_-enriched proteins had lower helix content (sec_helix_conf, mean 0.355 in (GT)_6_-SWCNT vs 0.442 in serum background, RBC = -0.230, FDR = 6.25×10^-15^) and higher sheet content (sec_sheet_conf, 0.258 vs 0.204, RBC = 0.206, FDR = 2.49×10^-12^), together with higher coil content (sec_coil_conf, 0.387 vs 0.354, RBC = 0.168, FDR = 1.35×10^-8^) and higher turn propensity (pp_ss_turn, 0.301 vs 0.287, RBC = 0.162, FDR = 1.92×10^-8^). Compositionally, (GT)_6_-bound proteins were enriched in glycine (aa_frac_G, 7.74 vs 6.84%, RBC = 0.146, FDR = 4.02×10^-7^), threonine (aa_frac_T, 5.67 vs 5.21%, RBC = 0.144, FDR = 4.85×10^-7^), cysteine (aa_frac_C, 2.67 vs 2.11%, RBC = 0.115, FDR = 5.98×10^-5^), and tyrosine (aa_frac_Y, 3.03 vs 2.78%, RBC = 0.104, FDR = 2.53×10^-4^), whereas leucine was depleted (aa_frac_L, 9.08 vs 9.73%, RBC = -0.171, FDR = 3.56×10^-9^). (GT)_6_-SWCNT proteins were also smaller (pp_length, 620 vs 755 aa, RBC = -0.155, FDR = 5.64×10^-8^; pp_mw, 68.6 vs 84.2 kDa, RBC = -0.160, FDR = 2.54×10^-8^), less basic (pp_isoelectric_point, 6.44 vs 6.75, RBC = -0.126, FDR = 9.93×10^-6^), and slightly more aromatic (pp_aromaticity, 0.0782 vs 0.0757, FDR = 8.76×10^-3^). Notably, (GT)_6_-enriched proteins also showed higher AlphaFold confidence, including higher AF_pLDDT_p10 (58.1 vs 55.0, FDR = 1.81×10^-4^) and lower AF_frac_pLDDT_lt50 (0.132 vs 0.174, FDR = 9.48×10^-6^), indicating that (GT)_6_-SWCNT preferentially recruited a relatively structured, non-helical, glycine/threonine/cysteine-enriched subset of serum proteins rather than simply favoring globally disordered proteins. Because a matched quantitative serum proteomics matrix was not available, we could not calculate a true abundance-normalized enrichment ratio of (GT)_6_-SWCNT relative to serum. We therefore performed a complementary quantitative analysis within the (GT)_6_-SWCNT corona itself using protein-group intensities, asking whether proteins that loaded more strongly into the (GT)_6_-SWCNT corona shared particular chemical properties (**Figure 7d**). Using Spearman rank correlation between feature values and mean (GT)_6_-SWCNT abundance, followed by Benjamini–Hochberg correction, the significant associations were higher cysteine fraction (aa_frac_C, ρ = 0.186, FDR = 1.29×10^-3^), higher tyrosine fraction (aa_frac_Y, ρ = 0.180, FDR = 1.29×10^-3^), higher aromaticity (pp_aromaticity, ρ = 0.164, FDR = 3.99×10^-3^), and higher tryptophan fraction (aa_frac_W, ρ = 0.151, FDR = 9.45×10^-3^), all associated with stronger (GT)_6_-SWCNT loading. In contrast, higher aspartate fraction (aa_frac_D, ρ = -0.144, FDR = 1.15×10^-2^) and higher alpha-helical propensity (pp_ss_helix, ρ = - 0.146, FDR = 1.15×10^-2^) were associated with lower (GT)_6_-SWCNT loading. Additional weaker but still significant associations included lower alanine fraction (aa_frac_A, ρ = -0.127, FDR = 3.33×10^-2^), higher serine fraction (aa_frac_S, ρ = 0.124, FDR = 3.62×10^-2^), and lower fraction of negatively charged residues (seq_frac_neg, ρ = -0.124, FDR = 4.21×10^-2^). We then applied the same framework to compare ALS and control coronas. At the protein level, we calculated a log2 fold-change for each (GT)_6_-SWCNT protein group from mean abundance in ALS relative to controls and tested whether this shift was associated with any chemical descriptor. This signal was weak: only serine fraction remained significant after correction (aa_frac_S, Spearman ρ = - 0.146, FDR = 0.0235), indicating that proteins relatively enriched in ALS tended to be slightly serine-poorer, while serine-rich proteins tended to be relatively more abundant in controls. All other single-feature associations failed FDR correction, and the multivariable model had essentially no predictive power (cross-validated R^2 ≈ 0), indicating that sequence chemistry alone did not strongly explain which individual proteins were relatively shifted between ALS and controls. However, when we examined the abundance-weighted chemical composition of the corona at the sample level, clearer group differences emerged. ALS-weighted coronas were shifted toward larger and heavier proteins (pp_length, FDR = 2.44×10^-4^; pp_mw, FDR = 2.44×10^-4^; Mass, FDR = 6.00×10^-4^), together with higher structural-confidence metrics (AF_pLDDT_p10, FDR = 2.44×10^-4^) and lower low-confidence fractions (AF_frac_pLDDT_lt50, FDR = 6.00×10^-4^).

Together, the mass-spectrometry results support a molecular basis for the spectral classification signal. ALS and control sera produced distinct SWCNT protein-corona compositions, with corrected enrichment for adaptive immune biology and nominal ALS-higher enrichment of inflammatory and complement-related proteins. In parallel, the global serum-background comparison showed that the (GT)_6_-SWCNT surface selects a chemically biased subset of serum proteins. These findings support the interpretation that the SWCNT sensor functions as an integrative transducer of disease-biased protein-corona composition rather than as a direct detector of a single analyte.

## 4. Discussion

In this study, we demonstrate a proof-of-concept of a novel integrative approach for blood-based ALS detection that combines multi-chirality time-resolved SWCNT near-infrared spectroscopy with deep learning. This time-resolved, multi-chirality ssDNA-SWCNT sensing platform, coupled with convolutional autoencoder analysis, can distinguish ALS from control serum and extract latent spectral features with clear biological relevance. In the current cohort of 39 individuals, comprising 20 ALS patients and 19 controls, serum exposure induced structured spectral evolution across the nanotube panel, and these responses supported derivation of latent variables that separated disease groups with strong performance. This approach represents a unicum in ALS detection because it does not rely on direct quantification of a predefined analyte; rather, it converts disease-dependent serum–nanotube interactions into a multivariate optical phenotype that can be decoded computationally. This distinction is important, because it places the platform closer to an integrative state-sensing strategy than to a conventional single-analyte assay.

A central finding is that the discriminative signal was not carried only by static spectral differences, but by the temporal evolution of the corona-dependent response across multiple chiralities **(Figure 3**). The 12-chirality array showed both shared and chirality-specific behavior: raw peak-intensity responses contained a strong common-mode component, but substantial non-redundant structure remained after common-mode removal. Temporal remodeling persisted after residualization, and the significant time × chirality interaction demonstrated that different chiralities followed distinct kinetic trajectories. This finding is consistent with prior evidence that protein–nanotube interactions depend on nanotube structure, including diameter, chiral angle, and electronic environment^15–17,47^. In our data, peak-intensity kinetics were governed primarily by tube diameter, whereas emission-center shifts showed a more complex dependence on both diameter and chiral angle. This dissociation suggests that intensity and wavelength shifts report complementary aspects of corona formation: intensity may reflect the extent or kinetics of adsorbed biomass accumulation, whereas emission shifts may reflect changes in the local dielectric environment and molecular organization at the nanotube surface. Thus, multi-chirality, multi-timepoint acquisition provides a physical basis for multiplexing beyond a single SWCNT optical channel.

The convolutional autoencoder architecture was specifically designed for this data modality and its architecture was selected on three grounds (**Figure 2**). First, individual chirality peaks present as local two-dimensional intensity distributions in excitation–emission space, and information content is carried by deformations of these local shapes (peak amplitude, broadening, and lateral shift). Convolutional filters suit detecting these patterns at the peak scale: each filter learns a 2D template that responds locally to a peak-like feature, and weight sharing reduces the parameter count by orders of magnitude relative to a fully connected encoder, which is essential at this sample size. We note that strict translation-equivariance does not hold for EEMs (chirality peaks occupy fixed, known coordinates rather than appearing at arbitrary positions) but the local-pattern argument and the parameter efficiency it confers do not depend on global equivariance. Convolutional architectures have been used analogously for multidimensional fluorescence spectral data, including excitation–emission matrices and three-dimensional fluorescence fingerprints, where positional information is meaningful and local-feature structure is preserved^52,53^. Second, pairing the classifier with a reconstruction objective converts every pixel of every EEM into a learning signal and acts as a strong unsupervised regularizer. The dual-objective loss ensures that the latent space preserves both the structural information of the EEMs (through reconstruction) and the disease-relevant patterns (through classification), preventing the model from collapsing into trivially discriminative but biologically uninterpretable features (**Figure 4**). We regard this as the principal architectural virtue of the convolutional autoencoder for this task. Third, encoding each timepoint through a shared encoder and aggregating via a learned temporal attention head keeps 0 h, 6 h, and 24 h in a common feature space, allowing the model to identify which phase of corona maturation carries the diagnostic signal, and yields interpretable per-subject temporal weights. The model assigned highest importance to the 6 h timepoint, corresponding to the intermediate stage of protein corona exchange when the Vroman effect is actively reshaping the corona composition^13,14^.

The demonstration of batch robustness addresses a critical challenge in biosensor development. The fact that proxy latent dimensions trained on Batch 1 achieve 97.4% accuracy on Batch 2 (SVM) indicates that the analytical latent dimension reconstruction pipeline, which uses the encoder’s own FC weights and attention mechanism to reconstruct latent values with zero free parameters, captures stable biological signal rather than batch-specific artifacts. Because it requires no additional fitting at the proxy-construction stage, this strategy reduces the risk of fitting batch-specific noise. This is critical for clinical translation, where reproducibility across different sensor preparations, time points, and laboratories is essential.

A second important conclusion is that the latent feature analysis suggests that the autoencoder captured information that was not readily recovered by conventional hand-engineered descriptors (**Figure 5**). Although the physical descriptor pipeline extracted peak amplitude, emission center, excitation center, FWHM, area, skewness, kurtosis, fit-quality metrics, chirality ratios, and inter-peak distances, these descriptors showed substantially weaker discrimination than the selected latent dimensions. Importantly, the best hand-engineered features were predominantly pairwise inter-chirality descriptors, indicating that ALS-associated information is distributed across relationships among nanotube species rather than localized to a single chirality peak. This supports the interpretation that the autoencoder did not simply rediscover one obvious optical descriptor; instead, it compressed distributed, nonlinear, multi-chirality spectral relationships into a small number of latent coordinates. Reconstruction-error and attribution analyses further support this interpretation, because important regions overlapped known SWCNT spectral coordinates rather than arbitrary image background.

At present, NfL remains the most established blood biomarker in ALS, with broad support as a marker of neuroaxonal injury, disease burden, and prognosis, but the field increasingly recognizes that NfL is not fully disease-specific and is unlikely to be sufficient on its own for all diagnostic and stratification purposes. Recent literature emphasizes that the future of ALS biomarker development lies in integrated, multimodal approaches combining neurodegeneration-linked markers with inflammatory, proteomic, and mechanistic signatures^50,51^. A recent Nature

Medicine study identified a plasma 33-protein biomarker panel that differentiated ALS from healthy and neurological disease controls across multiple cohorts and importantly showed that discriminative performance was retained even when NfL was excluded from the model^54^. In parallel, recent ALS biomarker reviews have similarly argued that the field is moving from reliance on single markers toward composite signatures that better reflect the biological heterogeneity of the disease. Our spectral biomarker aligns with this interpretation. Indeed, the strongest latent feature, dim477, was significantly associated with serum NfL, indicating that the SWCNT spectral phenotype covaries with a recognized marker of axonal injury (**Figure 6**).

However, this relationship was not reducible to a single optical descriptor or to a direct NfL readout. Single-chirality descriptors alone did not fully explain the dim477–NfL relationship, and the strongest shared associations involved inter-chirality relationships. Thus, dim477 is better interpreted as an NfL-associated multivariate corona signature that integrates axonal-injury-related biology with additional serum-corona information. Additionally, because NfL differs between ALS and control groups, future work should test whether the dim477–NfL relationship persists within ALS subjects alone or after adjustment for diagnosis. The Western blot experiments further support the biological plausibility of this interpretation. The (GT)_6_-SWCNT corona retained NfL, Tau, and NfH when incubated with serum protein extracts enriched with brain organoid homogenate, and Tau and NfH were detectable even in unspiked native serum-extracted corona samples. These findings do not prove that direct NfL adsorption drives dim477, nor that neurofilaments are the dominant causal species in native ALS serum coronas. They do, however, show that the nanotube corona can engage neurodegeneration-relevant proteins under serum-like conditions. This strengthens the interpretation that the spectral biomarker is connected to a biologically meaningful corona environment rather than to purely technical variation.

The mass spectrometry analysis provides critical biological validation (**Figure 7**). Compared with prior ALS blood proteomic studies, our results support a partially overlapping but more filtered immune-centered signature. After removal of likely contaminant, hemolysis-, platelet-, and coagulation-associated proteins, adaptive immune response remained the only significantly enriched pathway in our dataset (p = 1.03 × 10^-4^, FDR = 0.0354), whereas ALS-higher proteins were characterized by non-significant but convergent enrichment for inflammatory, complement-regulatory, and host-defense processes. This agrees with previous reports showing that ALS blood proteomics is enriched for immune and complement pathway alterations, including plasma studies reporting complement upregulation and complement cohort analyses showing higher circulating C4 and sC5b-9 in ALS^55,56^. It is also consistent with earlier serum proteome data identifying dysregulated acute-phase reactants and lipid-homeostasis proteins in ALS^57,58^. The main difference is that, in our filtered analysis, the strongest corrected signal localized to humoral/adaptive immune biology rather than complement itself, suggesting that once major pre-analytical confounders are removed, the ALS serum proteome may retain a broader immune remodeling pattern in which adaptive immune enrichment is statistically dominant, while innate inflammatory/complement activation remains present but less powered at the pathway level. The identification of PPARδ, collagen IV, and DMBT1 among enriched proteins points to metabolic, extracellular-matrix, and innate-immune processes with plausible relevance to neuroinflammation, neuromuscular-junction biology, and host-defense remodeling^59,60,61^.

Our (GT)_6_-SWCNT corona chemistry is broadly consistent with the selectivity principles proposed in previous studies^45,62^, but it also refines them in a way that appears specific to our filtered serum (GT)_6_-SWCNT system. Pinals et al. reported that, for ssDNA-SWCNT coronas, features linked to protein conformational adaptability, particularly glycine content and other non-secondary-structure-associated residues, contributed favorably to nanotube binding, whereas descriptors associated with internal structural stability, including leucine content, beta-sheet-associated amino acids, and GRAVY hydropathy, contributed unfavorably^45^. They further showed that proteins with higher glycine content preferentially bound to (GT)_6_ relative to (TAT)_4_, supporting glycine-mediated binding as a coating-specific interaction mode^62^. In our dataset, the strongest overlaps with Pinals et al. were the enrichment of glycine in the (GT)_6_-SWCNT corona (7.74% vs 6.84% in serum background; FDR = 4.02×10^-7^), the depletion of leucine (9.08% vs 9.73%; FDR = 3.56×10^-9^), and the positive contribution of glycine/flexibility-linked chemistry to (GT)_6_-SWCNT selectivity, all of which support the idea that (GT)_6_-SWCNT favors proteins with greater conformational adaptability rather than rigid hydrophobic packing. Compared with Pinals et al.^45^, our (GT)_6_-SWCNT corona retained the expected enrichment for flexible GT-associated sequence features but showed selectivity for structurally defined, non-helical, glycine/threonine/cysteine-enriched and leucine-depleted proteins, a difference that may reflect our serum pre-processing, buffer-exchange, and lower-SWCNT-concentration conditions.

In summary, the present work supports a formal model in which the ssDNA-SWCNT platform functions not as a direct one-analyte detector, but as an integrative transducer of disease-state protein corona dynamics. Relative to the current ALS biomarker landscape, where the field is increasingly converging on multi-marker plasma signatures rather than single-molecule readouts, this is a conceptually timely contribution. Relative to the SWCNT sensing literature, the study extends prior work on corona exchange and coating/chirality-dependent adsorption by showing that a fixed SWCNT chemistry, interrogated across multiple chirality and timepoints, can encode a disease-associated serum phenotype with both diagnostic and mechanistic relevance. The most compelling interpretation of the data is therefore not that the platform has already achieved a clinically mature ALS test, but that it establishes a strong proof of principle for a new class of neurodegenerative liquid biopsy in which complex biofluid composition is compressed into a learnable near-infrared spectral signature.

### Limitations

Several limitations should be noted. First, the cohort size was modest (N = 39). Although subject-level cross-validation, permutation testing, and same-subject batch-transfer experiments reduce concern that the results are purely technical artifacts, they do not replace independent validation in larger cohorts. Second, the current case-control design compared ALS patients with non-neurological controls. Clinical diagnostic utility will require testing against disease mimics and related neurodegenerative disorders, including frontotemporal dementia, progressive muscular atrophy, peripheral neuropathies, and inflammatory neuromuscular conditions. Third, Batch 2 tested technical reproducibility across experimental acquisition sessions using the same subjects, not generalization to new individuals. Fourth, mass spectrometry was performed on pooled ALS and control serum protein extracts, limiting individual-level inference about the relationship among corona composition, NfL, clinical phenotype, and spectral features.

Individual-level corona proteomics in larger cohorts will be necessary to identify the specific molecular drivers of the latent spectral signatures. Fifth, the relationship between dim477 and NfL may be partly driven by diagnostic group differences. Future studies should test whether this association persists within ALS subjects alone and after adjustment for disease severity, progression rate, age, sex, and other clinical covariates.

## Data Availability

All data produced in the present study are available upon reasonable request to the authors

---

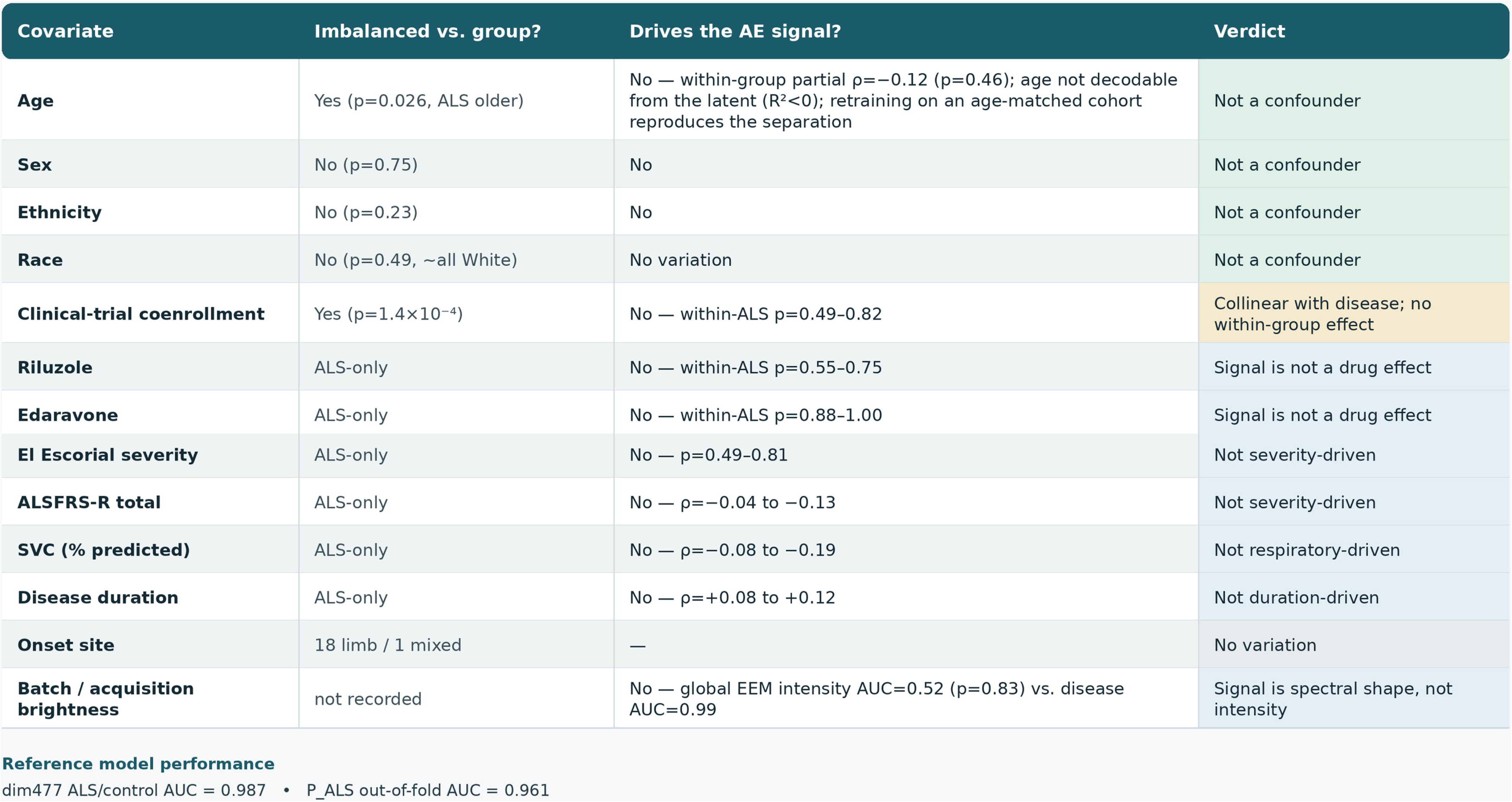

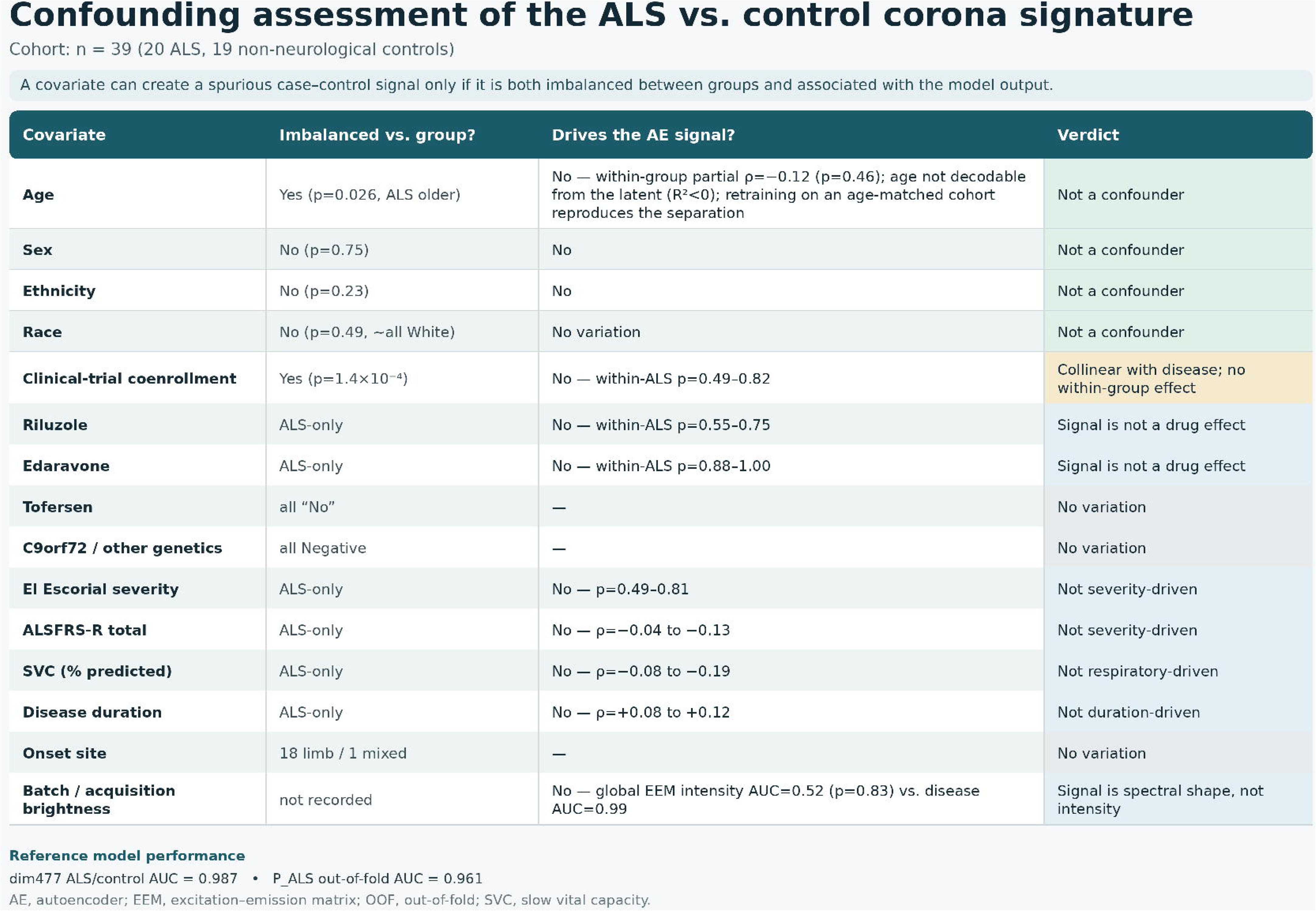

**Supplementary Table.**
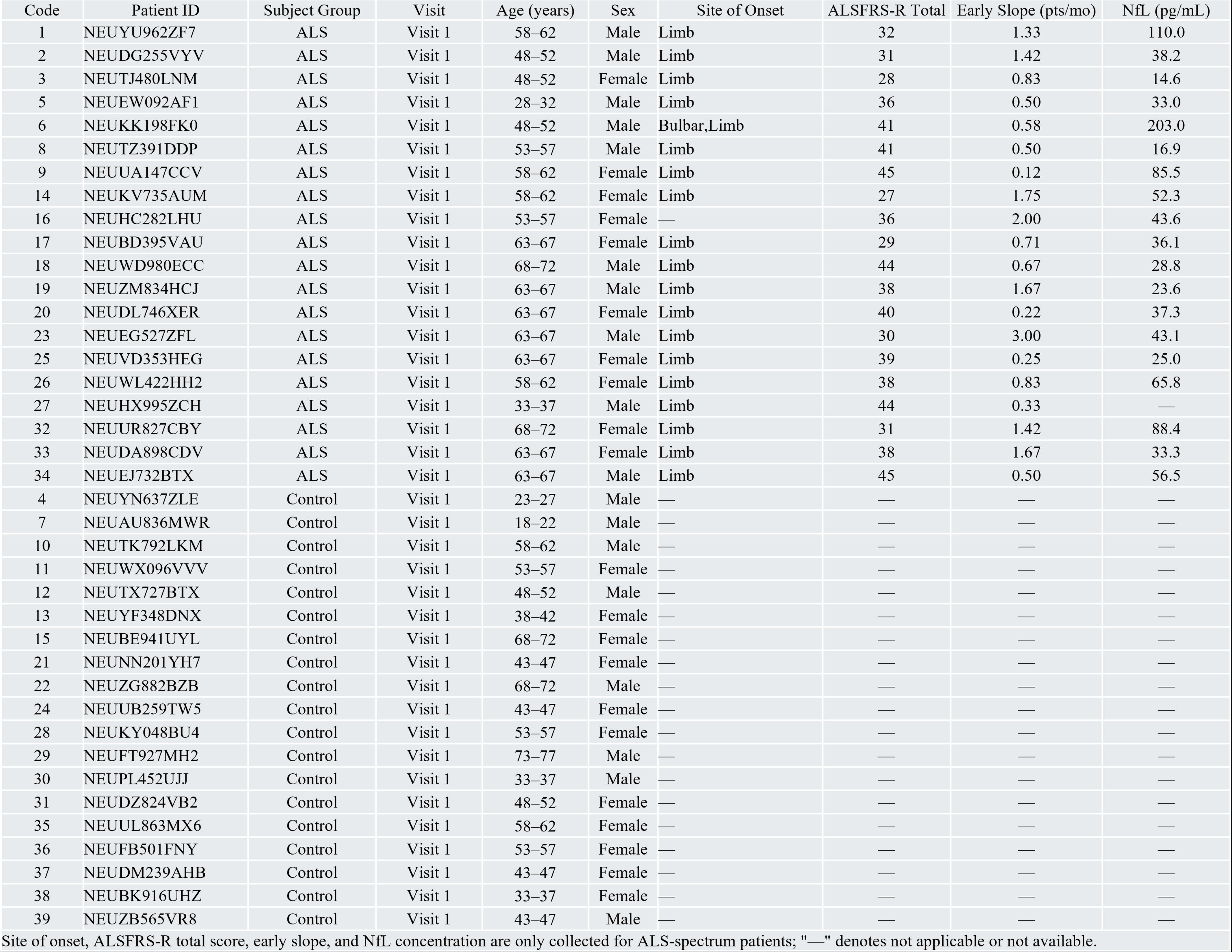
Patient cohort demographics and clinical characteristics.

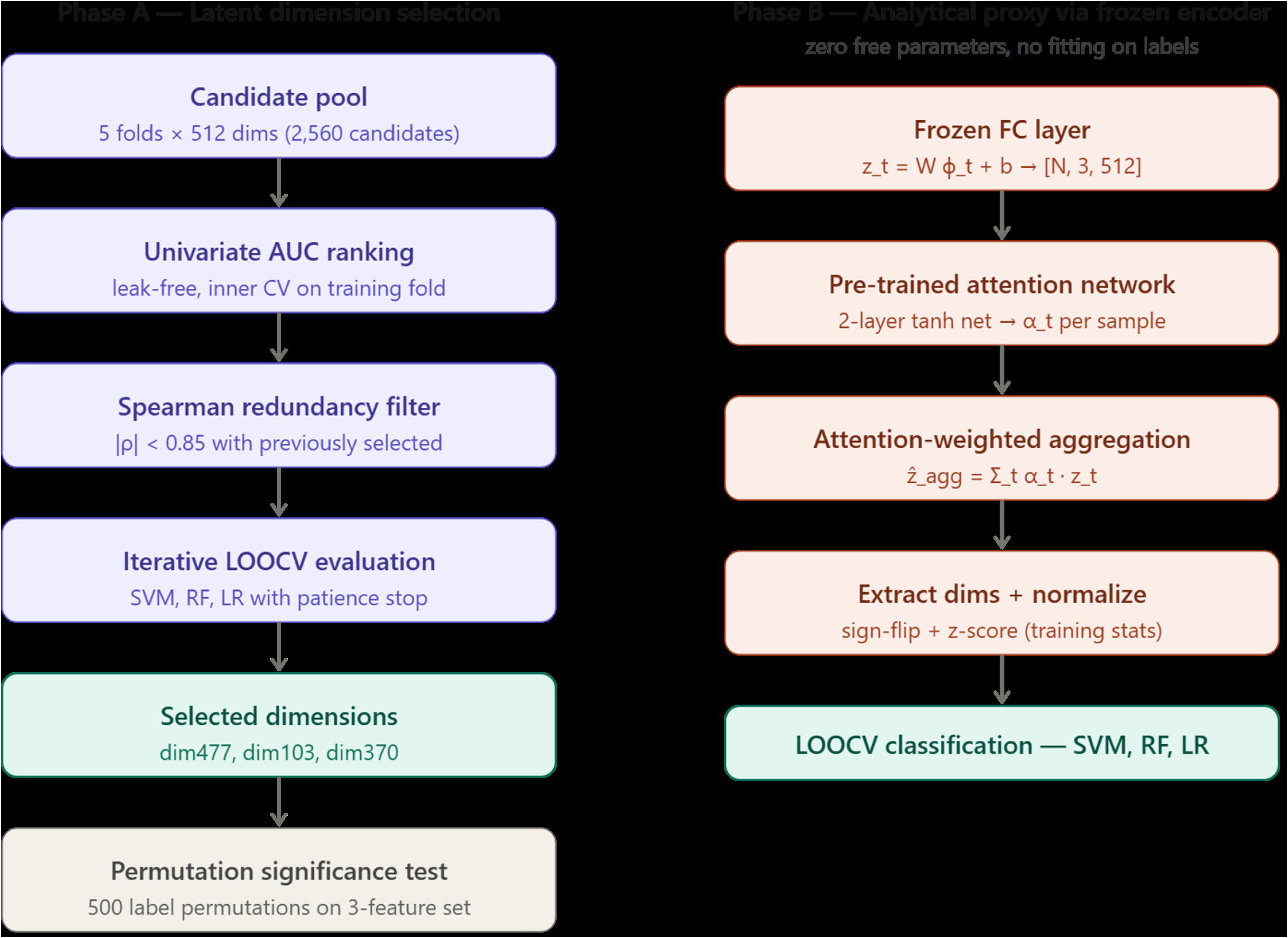

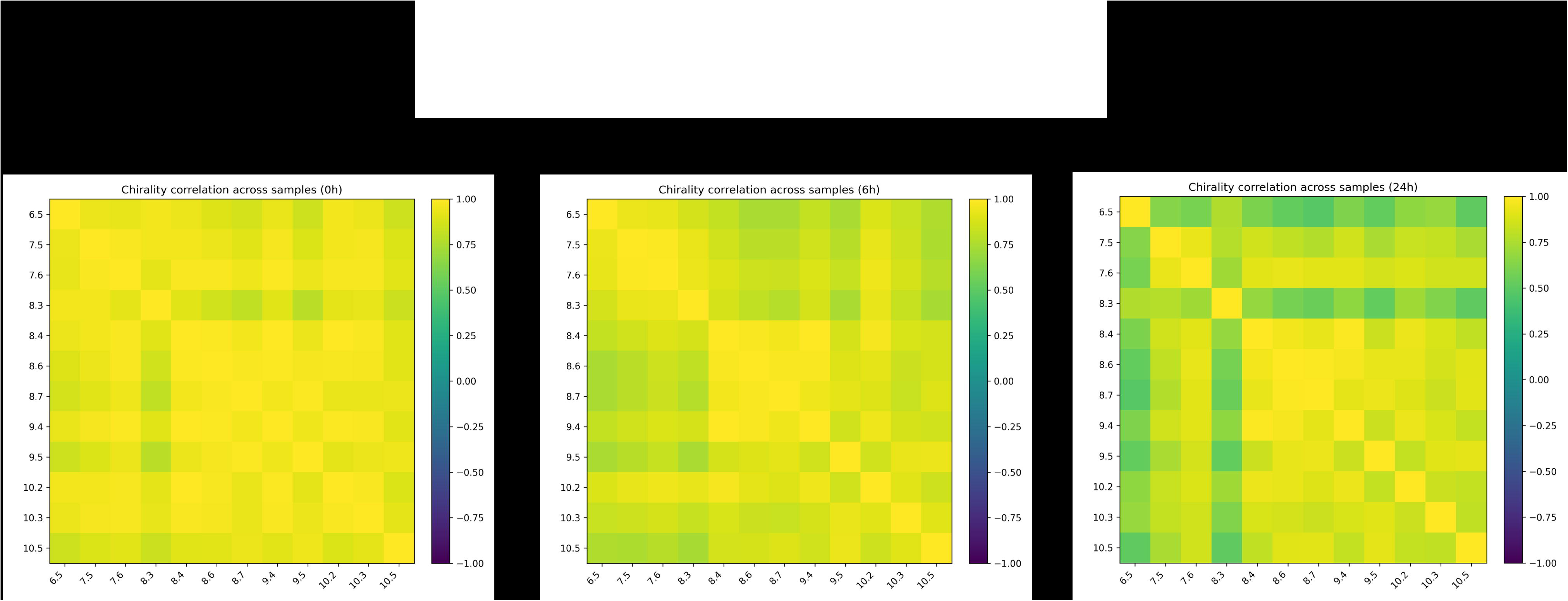

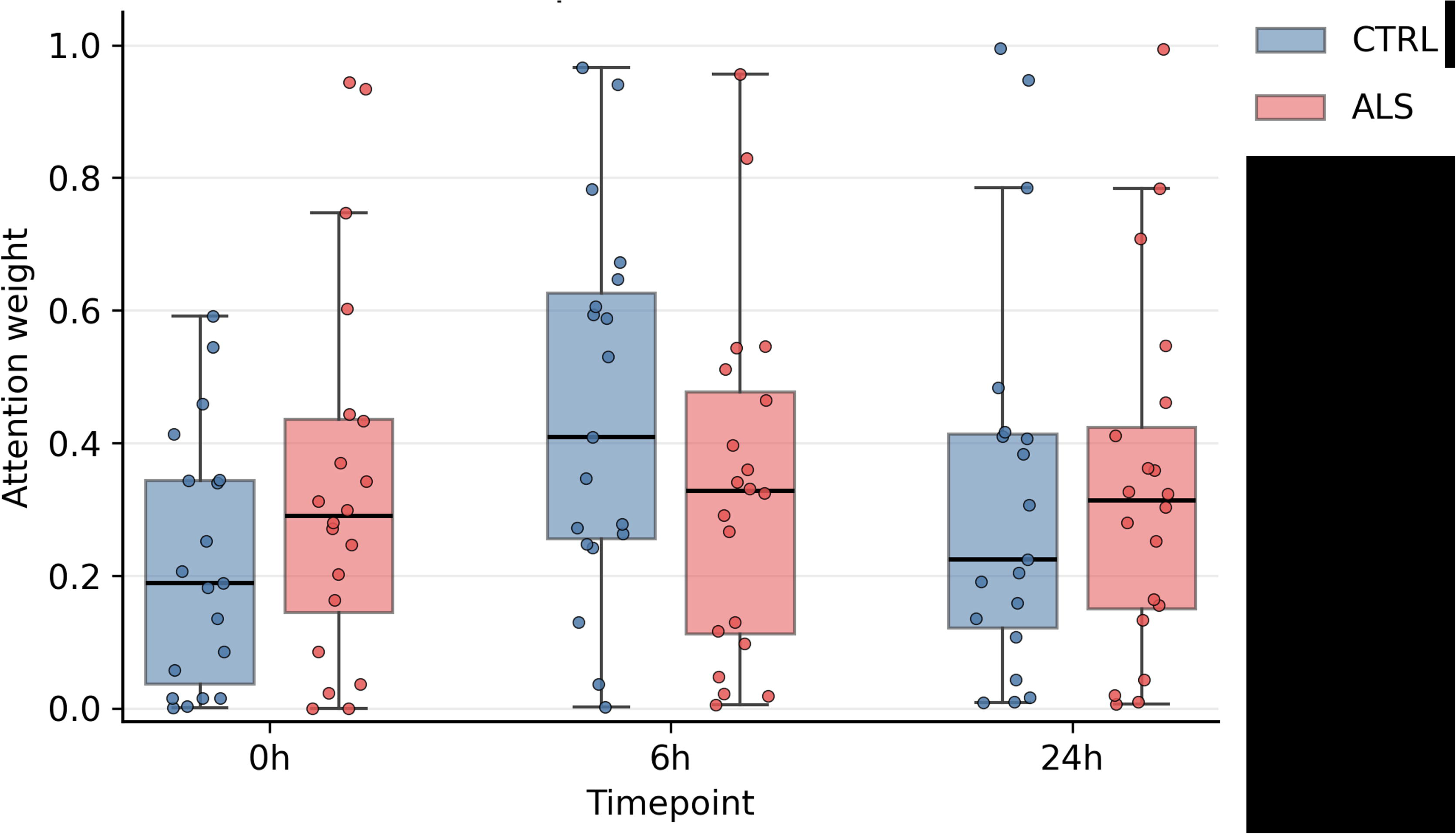

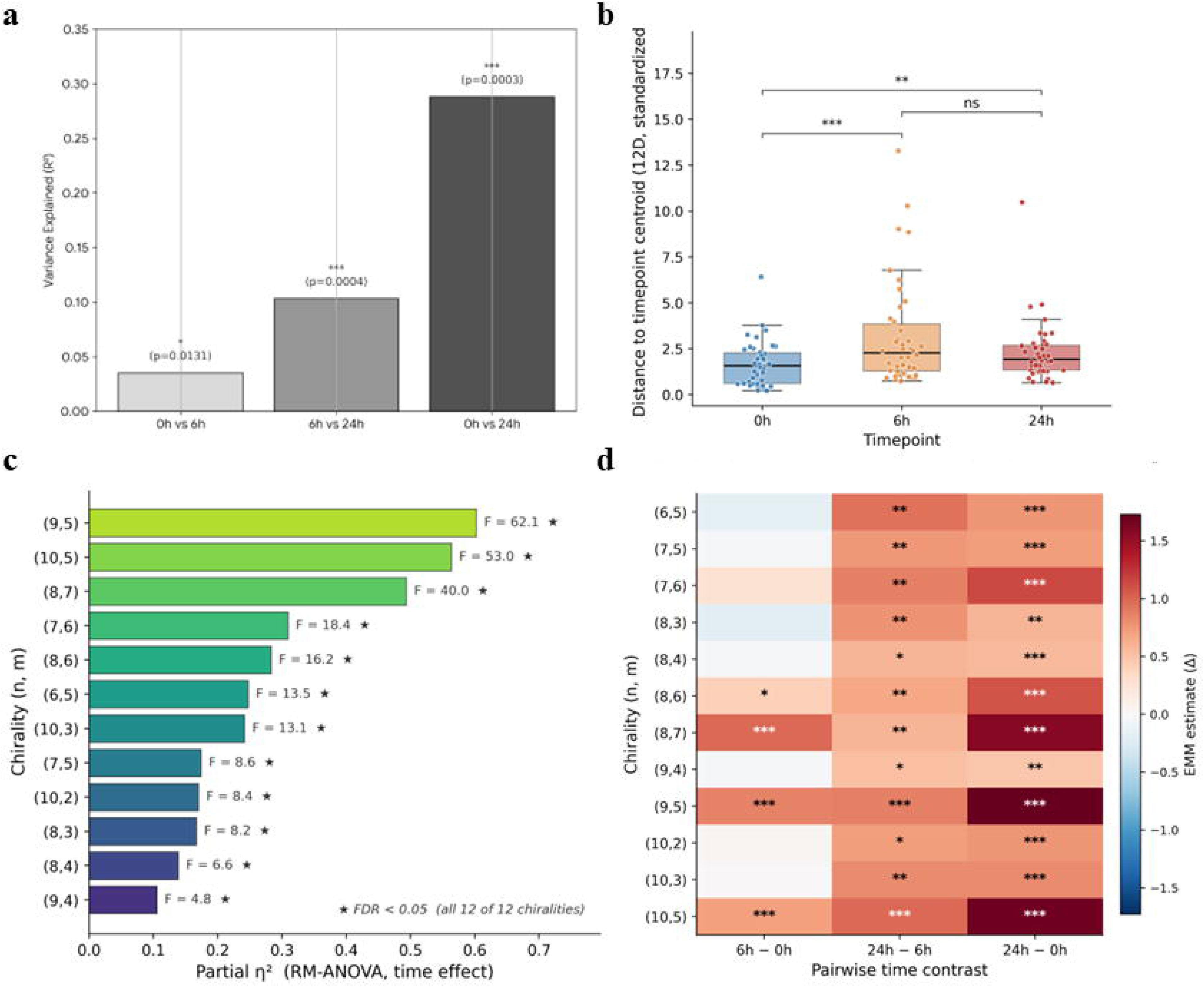

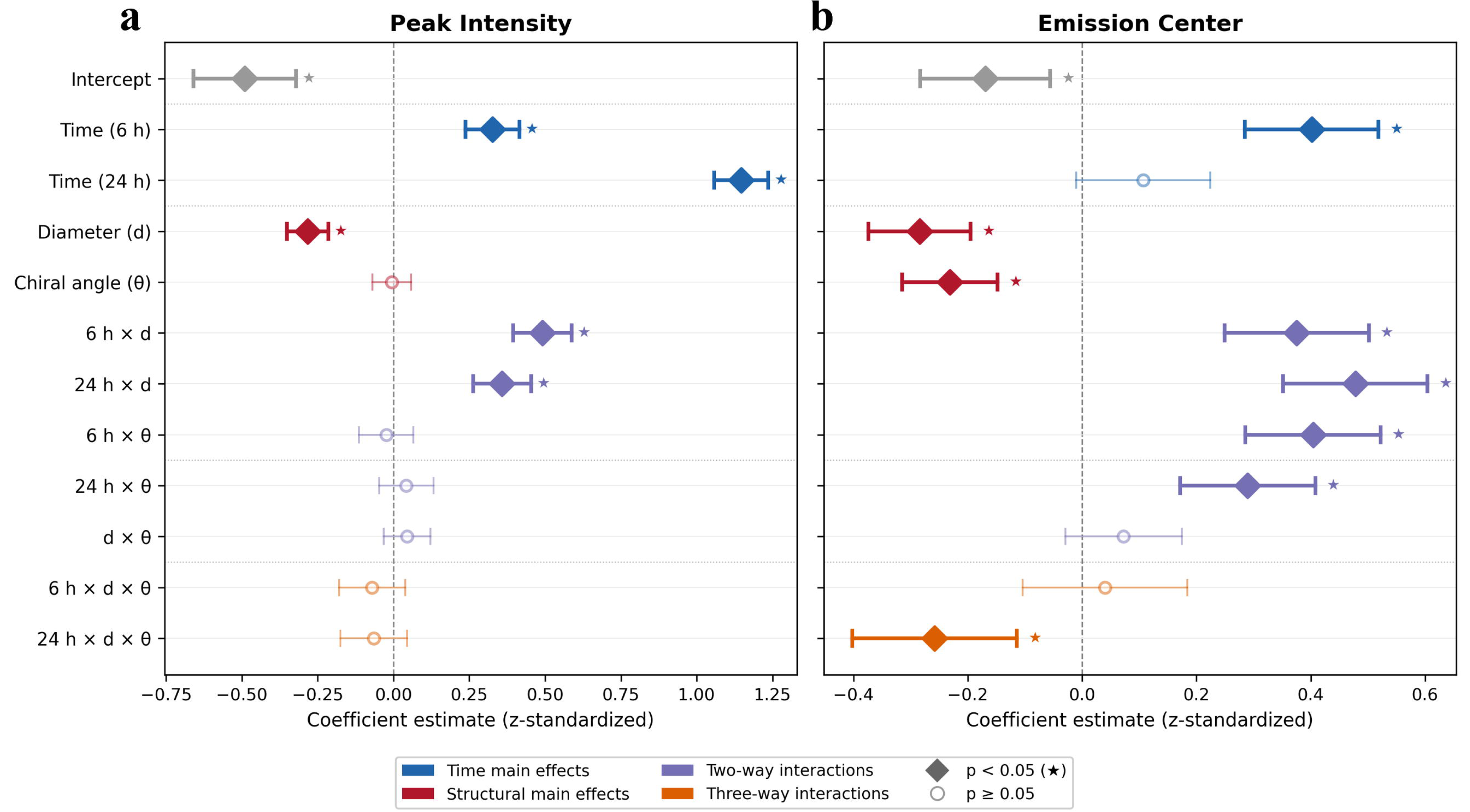

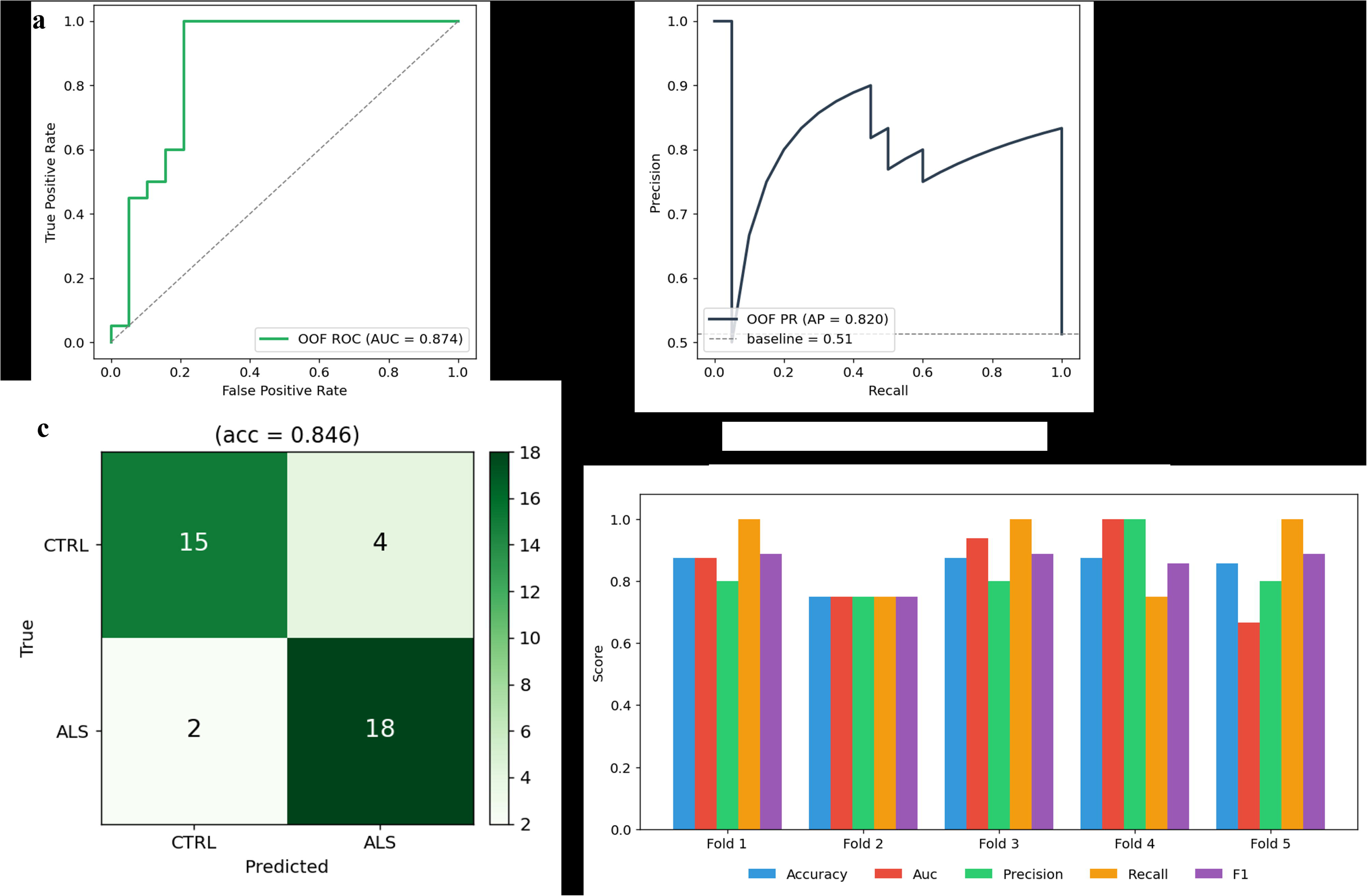

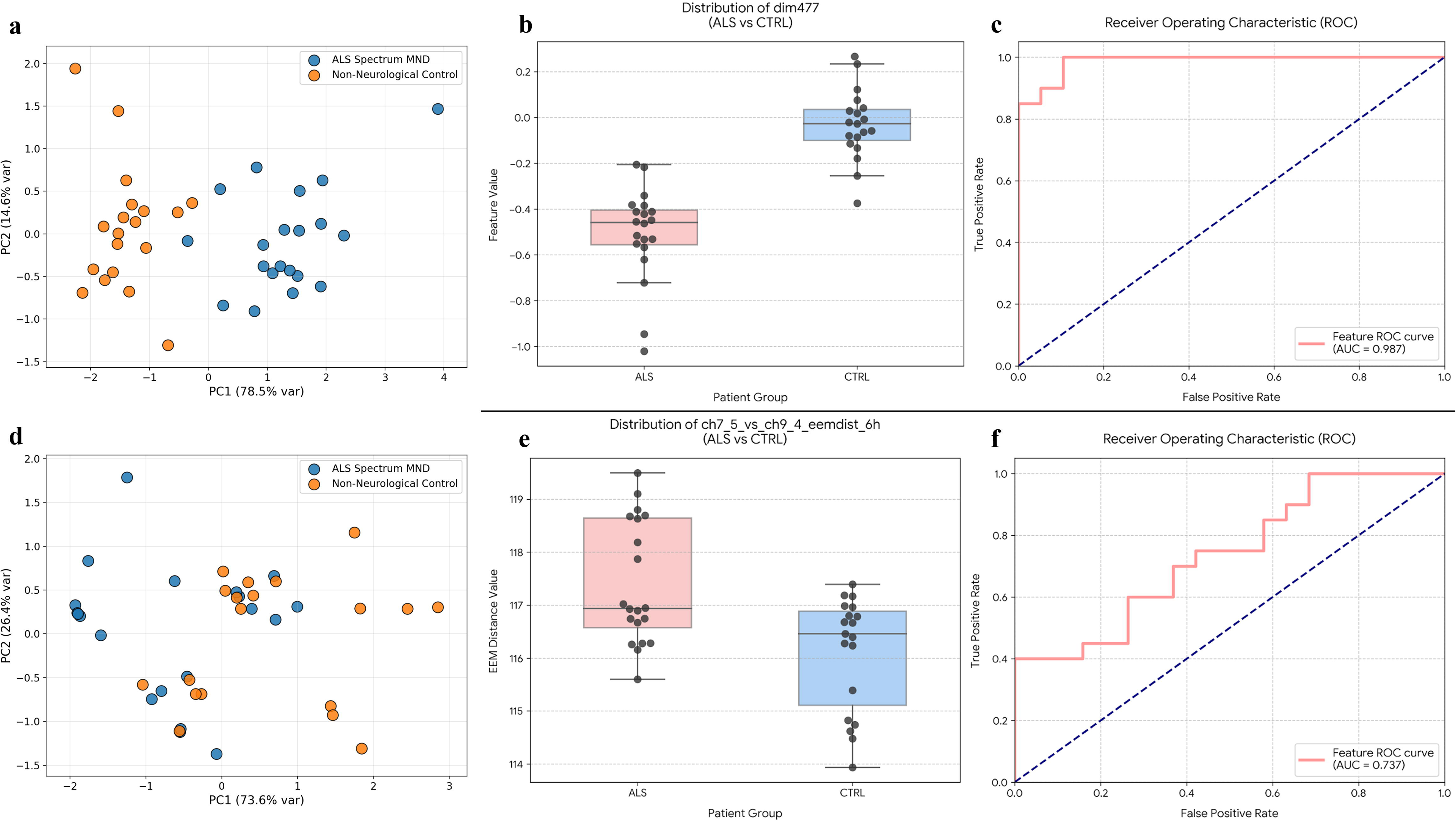

## References

1. Feldman, E. L. et al. Amyotrophic lateral sclerosis. Lancet 400, 1363–1380 (2022).

2. Arthur, K. C. et al. Projected increase in amyotrophic lateral sclerosis from 2015 to 2040. Nat. Commun. 7, 12408 (2016).

3. Mead, R. J., Shan, N., Reiser, H. J., Marshall, F. & Shaw, P. J. Amyotrophic lateral sclerosis: a neurodegenerative disorder poised for successful therapeutic translation. Nat. Rev. Drug Discov. 22, 185–212 (2023).

4. Verde, F. et al. Neurofilament light chain in serum for the diagnosis of amyotrophic lateral sclerosis. J. Neurol. Neurosurg. Psychiatry 90, 157–164 (2019).

5. Barschke, P., Oeckl, P., Steinacker, P., Ludolph, A. & Otto, M. Biomarkers for amyotrophic lateral sclerosis. Nat. Rev. Neurol. 18, 119–131 (2022).

6. Irwin, K. E. et al. A fluid biomarker reveals loss of TDP-43 splicing repression in presymptomatic ALS–FTD. Nat. Med. 30, 382–393 (2024).

7. Seddighi, S. et al. Mis-spliced transcripts generate de novo proteins in TDP-43-related ALS/FTD. Sci. Transl. Med. 16, eadg7162 (2024).

8. Bisker, G. et al. Protein-targeted corona phase molecular recognition. Nat. Commun. 7, 10241 (2016).

9. Nadeem, A. et al. Spectral fingerprinting of engineered nanomaterials for precision biosensing. ACS Nano 20, 3921–3943 (2026).

10. Kim, M. et al. Detection of ovarian cancer via the spectral fingerprinting of quantum-defect-modified carbon nanotubes in serum by machine learning. Nat. Biomed. Eng. 6, 267–275 (2022).

11. Kim, K. et al. Clinically accurate diagnosis of Alzheimer’s disease via multiplexed sensing of core biomarkers in human plasma. Nat. Commun. 11, 119 (2020).

12. FDA–NIH Biomarker Working Group. BEST (Biomarkers, EndpointS, and other Tools) Resource (Food and Drug Administration / National Institutes of Health, 2016).

13. Monopoli, M. P., Aberg, C., Salvati, A. & Dawson, K. A. Biomolecular coronas provide the biological identity of nanosized materials. Nat. Nanotechnol. 7, 779–786 (2012).

14. Tenzer, S. et al. Rapid formation of plasma protein corona critically affects nanoparticle pathophysiology. Nat. Nanotechnol. 8, 772–781 (2013).

15. Bachilo, S. M. et al. Structure-assigned optical spectra of single-walled carbon nanotubes. Science 298, 2361–2366 (2002).

16. Weisman, R. B. & Bachilo, S. M. Dependence of optical transition energies on structure for single-walled carbon nanotubes in aqueous suspension: an empirical Kataura plot. Nano Lett. 3, 1235–1238 (2003).

17. Saito, R., Dresselhaus, G. & Dresselhaus, M. S. Physical Properties of Carbon Nanotubes (Imperial College Press, London, 1998).

18. Brooks, B. R., Miller, R. G., Swash, M. & Munsat, T. L. El Escorial revisited: revised criteria for the diagnosis of amyotrophic lateral sclerosis. Amyotroph. Lateral Scler. Other Motor Neuron Disord. 1, 293–299 (2000).

19. Cedarbaum, J. M. et al. The ALSFRS-R: a revised ALS functional rating scale that incorporates assessments of respiratory function. J. Neurol. Sci. 169, 13–21 (1999).

20. Rissin, D. M. et al. Single-molecule enzyme-linked immunosorbent assay detects serum proteins at subfemtomolar concentrations. Nat. Biotechnol. 28, 595–599 (2010).

21. Roxbury, D. et al. Hyperspectral microscopy of near-infrared fluorescence enables 17-chirality carbon nanotube imaging. Sci. Rep. 5, 14167 (2015).

22. Bruderer, R. et al. Extending the limits of quantitative proteome profiling with data-independent acquisition and application to acetaminophen-treated three-dimensional liver microtissues. Mol. Cell. Proteomics 14, 1400–1410 (2015).

23. Li, H., Puopolo, T., Seeram, N. P., Liu, C. & Ma, H. Anti-ferroptotic effect of cannabidiol in human skin keratinocytes characterized by data-independent acquisition-based proteomics. J. Nat. Prod. 87, 1493–1499 (2024).

24. The UniProt Consortium. UniProt: the universal protein knowledgebase in 2025. Nucleic Acids Res. 53, D609–D617 (2025).

25. Benjamini, Y. & Hochberg, Y. Controlling the false discovery rate: a practical and powerful approach to multiple testing. J. R. Stat. Soc. B 57, 289–300 (1995).

26. Cock, P. J. A. et al. Biopython: freely available Python tools for computational molecular biology and bioinformatics. Bioinformatics 25, 1422–1423 (2009).

27. Jumper, J. et al. Highly accurate protein structure prediction with AlphaFold. Nature 596, 583–589 (2021).

28. Varadi, M. et al. AlphaFold Protein Structure Database: massively expanding the structural coverage of protein-sequence space with high-accuracy models. Nucleic Acids Res. 50, D439–D444 (2022).

29. Anderson, M. J. A new method for non-parametric multivariate analysis of variance. Austral Ecol. 26, 32–46 (2001).

30. Anderson, M. J., Ellingsen, K. E. & McArdle, B. H. Multivariate dispersion as a measure of beta diversity. Ecol. Lett. 9, 683–693 (2006).

31. Seabold, S. & Perktold, J. Statsmodels: econometric and statistical modeling with Python. Proc. 9th Python Sci. Conf. 92–96 (2010).

32. Wilks, S. S. The large-sample distribution of the likelihood ratio for testing composite hypotheses. Ann. Math. Stat. 9, 60–62 (1938).

33. Masci, J., Meier, U., Cireşan, D. & Schmidhuber, J. Stacked convolutional auto-encoders for hierarchical feature extraction. Lect. Notes Comput. Sci. 6791, 52–59 (2011).

34. Vaswani, A. et al. Attention is all you need. Adv. Neural Inf. Process. Syst. 30, 5998–6008 (2017).

35. Ioffe, S. & Szegedy, C. Batch normalization: accelerating deep network training by reducing internal covariate shift. Proc. 32nd Int. Conf. Mach. Learn. (ICML) 37, 448–456 (2015).

36. Nair, V. & Hinton, G. E. Rectified linear units improve restricted Boltzmann machines. Proc. 27th Int. Conf. Mach. Learn. (ICML), 807–814 (2010).

37. Srivastava, N., Hinton, G., Krizhevsky, A., Sutskever, I. & Salakhutdinov, R. Dropout: a simple way to prevent neural networks from overfitting. J. Mach. Learn. Res. 15, 1929–1958 (2014).

38. Paszke, A. et al. PyTorch: an imperative style, high-performance deep learning library. Adv. Neural Inf. Process. Syst. 32, 8024–8035 (2019).

39. Kingma, D. P. & Ba, J. Adam: a method for stochastic optimization. Proc. 3rd Int. Conf. Learn. Represent. (ICLR) (2015).

40. Simonyan, K., Vedaldi, A. & Zisserman, A. Deep inside convolutional networks: visualising image classification models and saliency maps. Proc. Int. Conf. Learn. Represent. Workshop (2014).

41. Sundararajan, M., Taly, A. & Yan, Q. Axiomatic attribution for deep networks. Proc. 34th Int. Conf. Mach. Learn. (ICML) 70, 3319–3328 (2017).

42. Alain, G. & Bengio, Y. Understanding intermediate layers using linear classifier probes. Proc. Int. Conf. Learn. Represent. Workshop (2017).

43. Pedregosa, F. et al. Scikit-learn: machine learning in Python. J. Mach. Learn. Res. 12, 2825–2830 (2011).

44. Sirtori, Riccardo et al. A tabletop blast device for the study of the long-term consequences of traumatic brain injury on brain organoids. Cell Reports Methods vol. 5,11 (2025).

45. Pinals, R. L., et al. Quantitative protein corona composition and dynamics on carbon nanotubes in biological environments. Angew. Chem. Int. Ed. 59, 23668–23677 (2020).

46. Pinals, R. L., Yang, D., Lui, A., Cao, W. & Landry, M. P. Corona exchange dynamics on carbon nanotubes by multiplexed fluorescence monitoring. J. Am. Chem. Soc. 142, 1254–1264 (2020).

47. Salem, D. P. et al. Chirality dependent corona phase molecular recognition of DNA-wrapped carbon nanotubes. Carbon 97, 147–153 (2016).

48. Larsen, B. A. et al. Effect of solvent polarity and electrophilicity on quantum yields and solvatochromic shifts of single-walled carbon nanotube photoluminescence. J. Am. Chem. Soc. 134, 12485–12491 (2012).

49. Nakagawa, S., Johnson, P. C. D. & Schielzeth, H. The coefficient of determination R² and intra-class correlation coefficient from generalized linear mixed-effects models revisited and expanded. J. R. Soc. Interface 14, 20170213 (2017).

50. Thompson, A. G. et al. Multicentre appraisal of amyotrophic lateral sclerosis biofluid biomarkers shows primacy of blood neurofilament light chain. Brain Commun. 4, fcac029 (2022).

51. McMackin, R., Bede, P., Pender, N., Hardiman, O. & Nasseroleslami, B. Biomarkers in amyotrophic lateral sclerosis: current status and future prospects. Nat. Rev. Neurol. 19, 754–768 (2023).

52. Rutherford, J. W. et al. Excitation emission matrix fluorescence spectroscopy for combustion generated particulate matter source identification. Atmos. Environ. 220, 117065 (2020).

53. Ju, L. et al. Deep learning-assisted three-dimensional fluorescence difference spectroscopy for identification and semiquantification of illicit drugs in biofluids. Anal. Chem. 91, 9343–9347 (2019).

54. Chia, R., Moaddel, R., Kwan, J. Y. et al. A plasma proteomics-based candidate biomarker panel predictive of amyotrophic lateral sclerosis. Nat. Med. 31, 3440–3450 (2025).

55. Mantovani, S. et al. Elevation of the terminal complement activation products C5a and C5b-9 in ALS patient blood. J. Neuroimmunol. 276, 213–218 (2014).

56. Kjældgaard, A. L. et al. Complement profiles in patients with amyotrophic lateral sclerosis: a prospective observational cohort study. J. Inflamm. Res. 14, 1043–1053 (2021).

57. Mariosa, D. et al. Blood biomarkers of carbohydrate, lipid, and apolipoprotein metabolisms and risk of amyotrophic lateral sclerosis: a more than 20-year follow-up of the Swedish AMORIS cohort. Ann. Neurol. 81, 718–728 (2017).

58. Phan, K. et al. Multiple pathways of lipid dysregulation in amyotrophic lateral sclerosis. Brain Commun. 5, fcac356 (2023).

59. Iannotti, F. A. & Vitale, R. M. Recent insights on the role of PPAR-β/δ in neuroinflammation and neurodegeneration, and its potential target for therapy. NeuroMolecular Med. 23, 28–41 (2021).

60. Sanes, J. R., Engvall, E., Butkowski, R. & Hunter, D. D. Molecular heterogeneity of basal laminae: isoforms of laminin and collagen IV at the neuromuscular junction and elsewhere. J. Cell Biol. 111, 1685–1699 (1990).

61. Madsen, J., Mollenhauer, J. & Holmskov, U. Gp-340/DMBT1 in mucosal innate immunity. Innate Immun. 16, 160–167 (2010).

62. Ouassil, N., Pinals, R. L., Del Bonis-O’Donnell, J. T., Wang, J. W. & Landry, M. P. Supervised learning model predicts protein adsorption to carbon nanotubes. Sci. Adv. 8, eabm0898 (2022).

