## Supplementary Figures for "Multiplexed temporal SWCNT biosensor combined with convolutional autoencoding identifies ALS-associated serum protein corona signatures"

### Supplementary Material

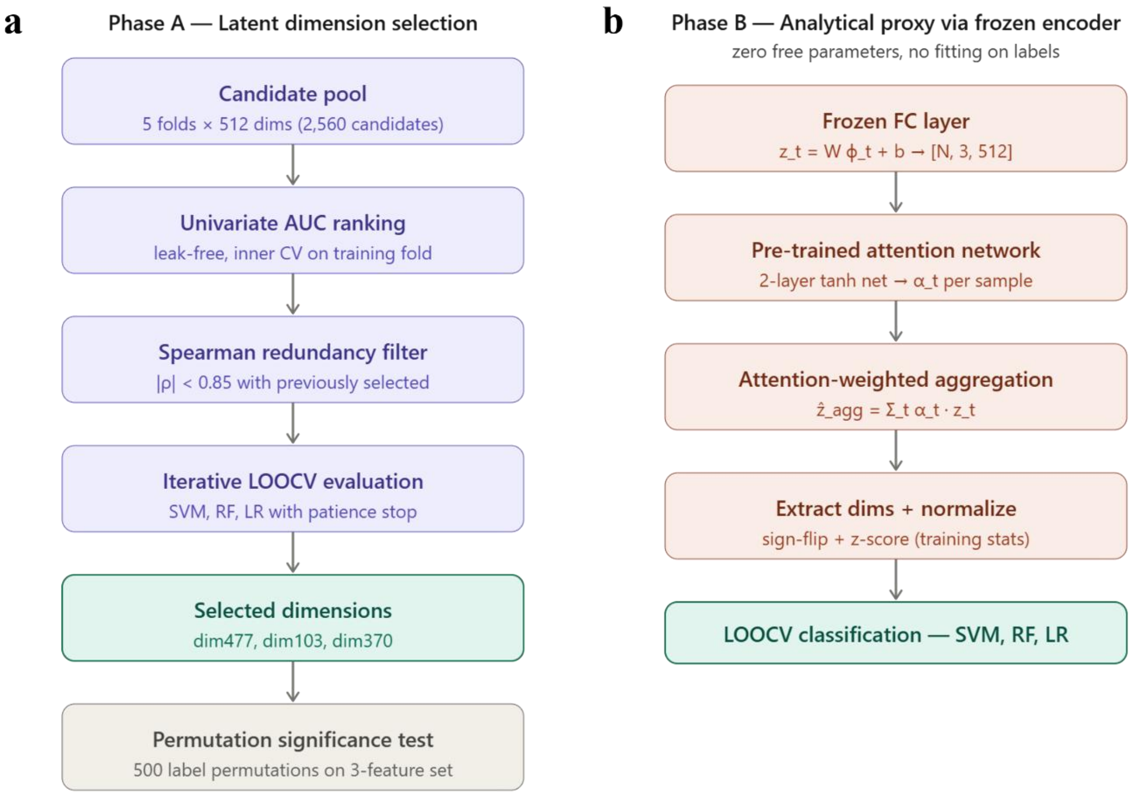


***Supplementary Figure 1.*** Latent dimension selection **(a)** and analytical proxy reconstruction **(b)**.

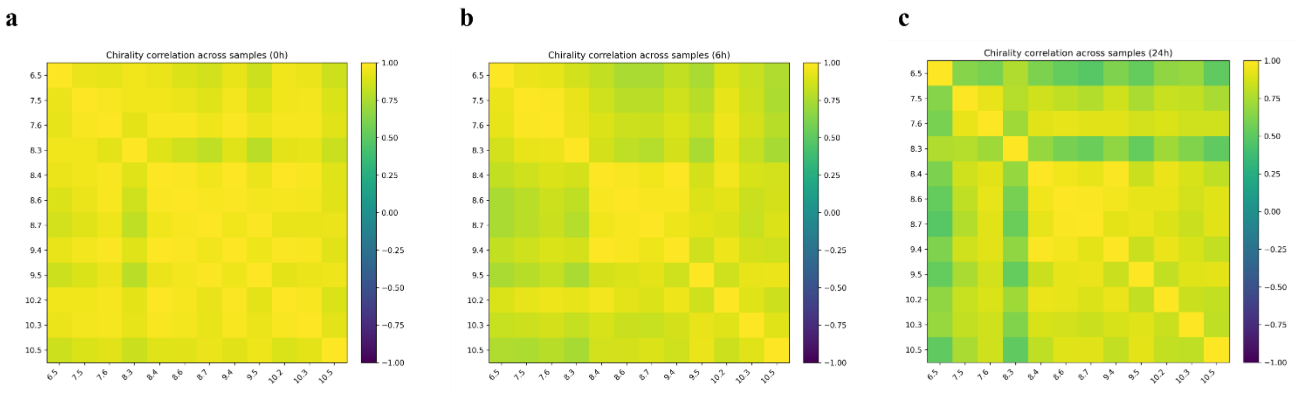


***Supplementary Figure 2. (a–c)*** Inter-chirality Pearson correlation matrices at 0 h, 6 h, and 24 h, showing progressive decorrelation as protein corona matures.

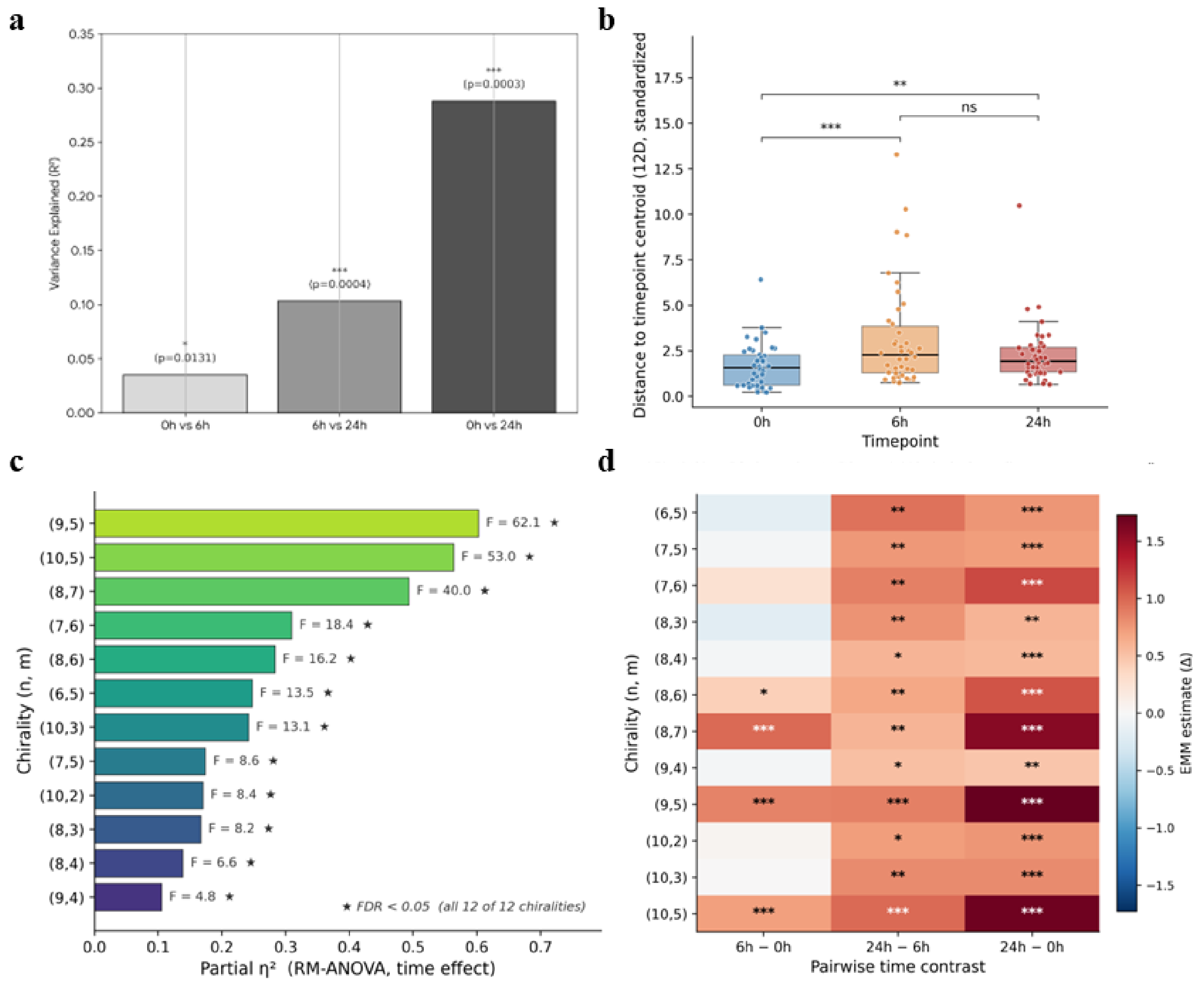


***Supplementary Figure 3.*** ***(a)*** Pairwise PERMANOVA on the 12-chirality EEM feature vectors across timepoints. Bars show the variance explained ($R^2$) by timepoint identity for each pairwise comparison (0 h vs. 6 h, 6 h vs. 24 h, 0 h vs. 24 h); Holm-corrected p-values from 9,999 permutations are annotated above each bar. The progressive increase in $R^2$ (0.035 → 0.10 → 0.29) indicates that the spectral separation between timepoints grows monotonically with corona maturation. **(b)** PERMDISP analysis of multivariate dispersion across time-points. Each point is the Euclidean distance from one sample's standardized 12-chirality feature vector to the centroid of its own timepoint group; boxes show median and interquartile range. Dispersion is lowest at 0 h, expands significantly by 6 h, and partially contracts by 24 h while remaining elevated above baseline. Brackets indicate pairwise paired Wilcoxon tests (Holm-corrected): *** $p < 0.001$, ** $p < 0.01$, ns = not significant; global Friedman $\chi^2 = 11.76$, $p = 0.003$. **(c)** Per-chirality repeated-measures ANOVA for the time effect. Bars show partial $\eta^2$ (effect size) for each of the 12 chiralities, sorted by magnitude; F-statistics are annotated next to each bar. All 12 chiralities exhibit FDR-significant temporal changes (★, Benjamini–Hochberg $q < 0.05$ across the chirality family), but with effect sizes spanning more than a fivefold range — from (9,5), (10,5), and (8,7)

at the high end (partial $\eta^2 > 0.49$) to (9,4) at the low end (partial $\eta^2 = 0.11$), indicating that chiralities evolve along independent kinetic trajectories. **(d)** Post-hoc estimated marginal mean (EMM) contrasts from the linear mixed model with sample as random effect, displayed as a chirality × time-contrast heatmap. Cell color encodes the estimated marginal difference (Δ) between paired timepoints; asterisks mark FDR-corrected significance (* $q < 0.05$, ** $q < 0.01$, *** $q < 0.001$). 28 of 36 contrasts (78%) are FDR-significant, and the 24 h − 0 h contrast is significant for all 12 chiralities; the LMM time × chirality interaction is highly significant ($p < 0.0001$), confirming chirality-resolved kinetic decorrelation as the basis for spectral multiplexing.

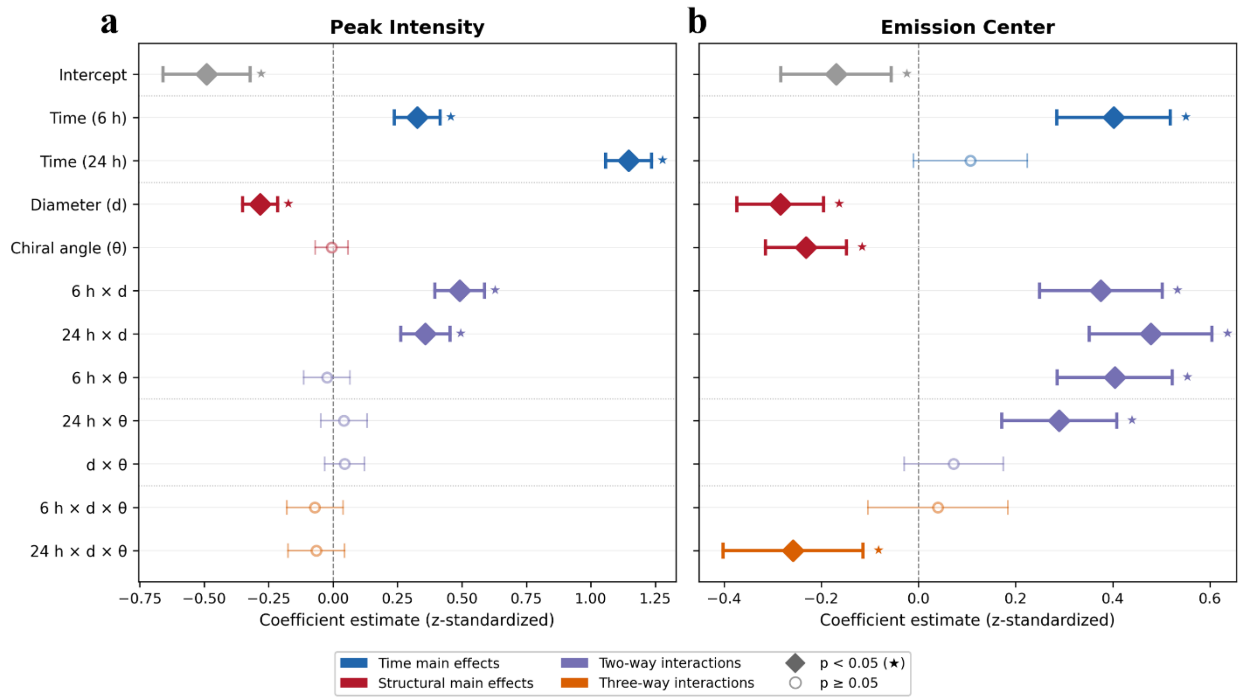


***Supplementary Figure 4.*** **Linear mixed-model coefficients linking SWCNT structural parameters to corona-driven spectral changes.** Forest plots of fixed-effect estimates from linear mixed models predicting two physical descriptors of the SWCNT emission, peak intensity **(a)** and emission center wavelength **(b)**, as a function of time (6 h and 24 h vs. 0 h reference), tube diameter (d), chiral angle (θ), and all two- and three-way interactions. Sample identity was modeled as a random intercept to account for repeated measurements across timepoints. All predictors and outcomes were z-standardized so that coefficients are directly comparable in magnitude across panels and terms. Points show the point estimate, horizontal bars indicate 95% confidence intervals, and filled diamonds with stars (*) denote terms with $p < 0.05$; open circles indicate non-significant terms ($p \geq 0.05$). Colors group the terms into time main effects (blue), structural main effects (red), two-way interactions (purple), and the three-way interaction (orange). **(a)** For peak intensity, both time main effects are positive and increase monotonically (24 h ≫ 6 h), diameter exerts a negative main effect (smaller-diameter chiralities brighten more), and the time × diameter interactions are positive and significant at both timepoints, indicating that the diameter-dependent intensity gain grows with corona maturation. Chiral angle and all interactions involving θ, including the three-way time × d × θ term, are not significant for peak intensity, supporting an additive model in which tube diameter is the dominant structural predictor of intensity kinetics. **(b)** For emission center, the pattern is more complex: both diameter and chiral angle contribute significant negative main effects, the time × diameter and time × chiral-angle interactions are significant at both 6 h and 24 h, and the three-way interaction reaches significance at 24 h (24 h × d × θ). This indicates that the magnitude and direction of the corona-induced solvatochromic shift depend jointly on tube geometry, including the interplay between diameter and chiral angle, and that this geometric coupling becomes most pronounced once the hard corona is fully formed.

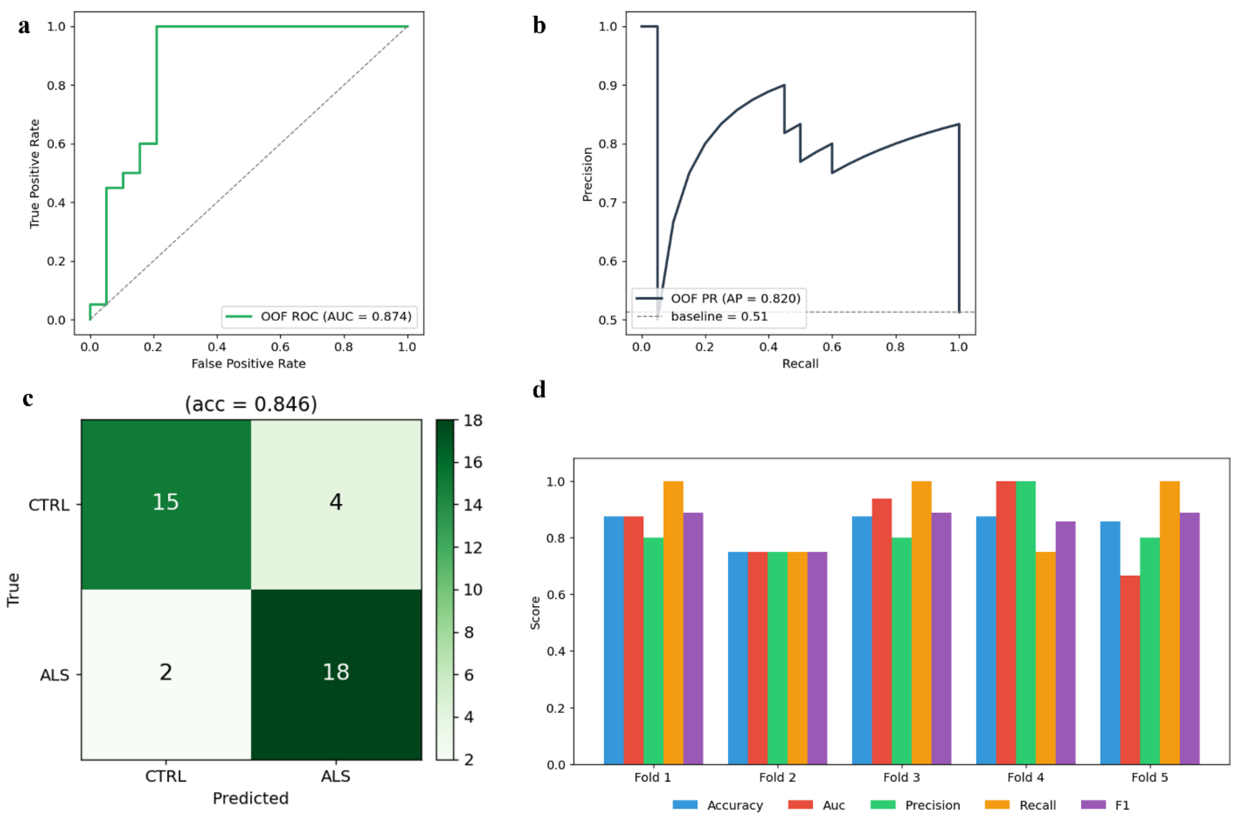


***Supplementary Figure 5*. Cross-validated performance of the convolutional autoencoder–classifier on EEM fluorescence spectra**. Dual-objective model (image reconstruction + binary ALS-vs-CTRL classification with attention over the 0 h/6 h/24 h timepoints) evaluated by subject-level 5-fold stratified cross-validation (n = 39; 20 ALS, 19 CTRL).

**(a)** ROC curve (OOF). Pooled out-of-fold receiver operating characteristic; area under the curve AUC = 0.87. Dashed line = chance. **(b)** Precision–recall curve (OOF). Average precision AP = 0.82; dashed line = positive-class prevalence (0.51). **(c)** Confusion matrix (OOF). Counts at a 0.5 probability threshold (overall accuracy = 0.85). Controls: 15/19 correct (4 false positives); ALS: 18/20 correct (2 false negatives). **(d)** Per-fold validation metrics. Accuracy, AUC, precision, recall and F1 for each of the 5 held-out folds, illustrating fold-to-fold variability typical of a small cohort (fold AUCs 0.75–1.00).

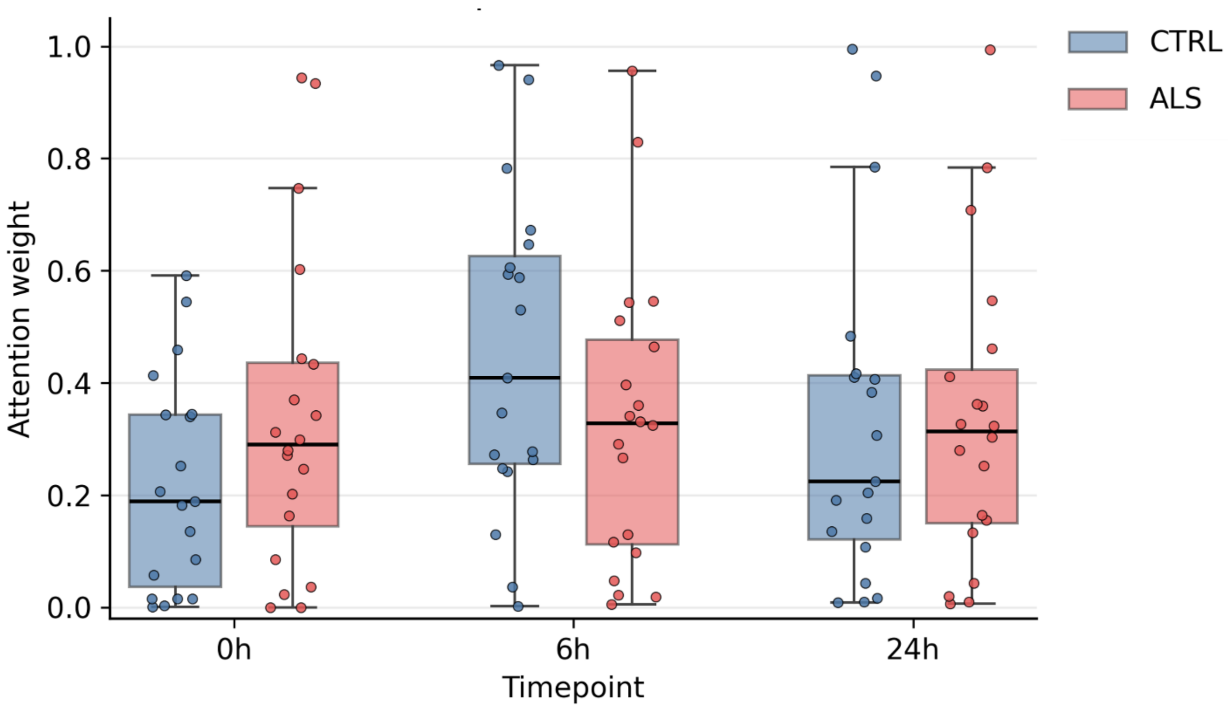


***Supplementary Figure 6.*** **Subject-level temporal attention weights from the convolutional autoencoder classifier** (5-fold cross-validation, held-out predictions; n = 39 subjects: 20 ALS, 19 CTRL). Each dot represents one subject's attention weight at the indicated timepoint, taken from the fold in which that subject was held out for validation.

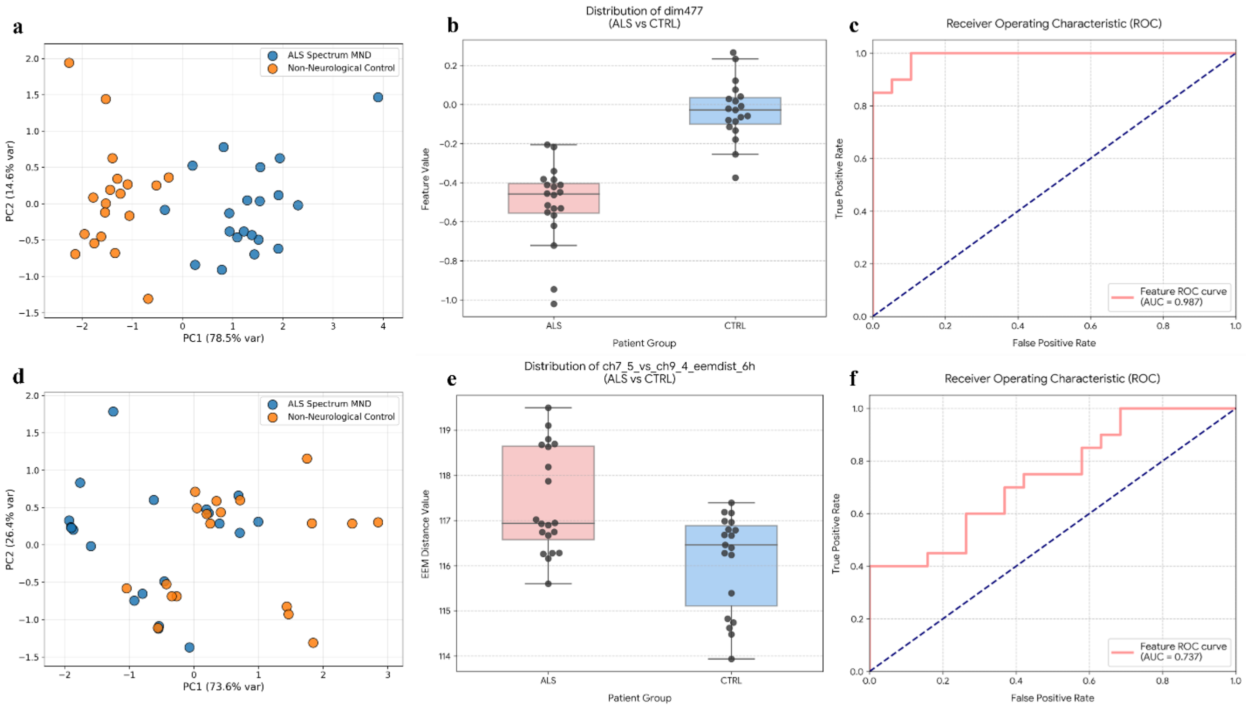


**Supplementary Figure 7. Head-to-head comparison of learned (autoencoder) vs. hand-crafted physical descriptors for ALS classification**. **(a–c)** Top-ranked autoencoder latent features. **(d–f)** Top-ranked hand-crafted physical descriptors. In both cases features were selected by the leak-free univariate AUC ranking with stability filtering across subject-level 5-fold cross-validation, as described in the Methods. **(a)** Principal-component projection of the samples in the space of the top-ranked autoencoder latent features (selection procedure described in the Methods). PC1 (78.5% of variance) cleanly separates ALS-spectrum motor neuron disease cases (blue) from non-neurological controls (orange), indicating that the latent feature space captures a dominant axis of group structure. **(b)** Distribution of the single most discriminative latent feature (dim477, drawn from fold 3 of the best-performing model) in ALS versus control sera. The two groups occupy nearly non-overlapping regimes (ALS centered around −0.45, controls near 0), with a clear shift in median and a well-separated interquartile range. **(c)** Receiver operating characteristic curve for the same latent feature (dim477) used as a univariate classifier; area under the curve (AUC) = 0.987, demonstrating near-perfect ranking of ALS vs. control sera by this single learned feature. **(d)** PCA projection in the space of the top-ranked hand-crafted physical descriptors (cross-chirality EEM-distance, peak, and shape descriptors; selection procedure described in the Methods). PC1 (73.6% of variance) does not separate ALS from controls, and the two groups overlap substantially across both principal components. **(e)** Distribution of the single best-performing hand-crafted feature, the EEM distance between the (7,5) and (9,4) chirality peaks at 6 h (ch7_5_vs_ch9_4_eemdist_6h), in ALS versus control sera. The means differ in the expected direction, but the distributions overlap heavily. **(f)** ROC curve for the same hand-crafted feature used as a univariate classifier; AUC = 0.737, well below the

0.987 achieved by the top latent feature in panel **(c)** and consistent with the visibly weaker class separation in panels **(d)** and **(e).**

| Supplementary Table — Patient cohort demographics and clinical characteristics | | | | | | | | | |
|---|---|---|---|---|---|---|---|---|---|
| Code | Patient ID | Subject Group | Visit | Age (years) | Sex | Site of Onset | ALSFRS-R Total | Early Slope (pts/mo) | NfL (pg/mL) |
| 1 | NEUYU962ZF7 | ALS | Visit 1 | 58–62 | Male | Limb | 32 | 1.33 | 110.0 |
| 2 | NEUDG255VYV | ALS | Visit 1 | 48–52 | Male | Limb | 31 | 1.42 | 38.2 |
| 3 | NEUTJ480LNM | ALS | Visit 1 | 48–52 | Female | Limb | 28 | 0.83 | 14.6 |
| 5 | NEUEW092AF1 | ALS | Visit 1 | 28–32 | Male | Limb | 36 | 0.50 | 33.0 |
| 6 | NEUKK198FK0 | ALS | Visit 1 | 48–52 | Male | Bulbar,Limb | 41 | 0.58 | 203.0 |
| 8 | NEUTZ391DDP | ALS | Visit 1 | 53–57 | Male | Limb | 41 | 0.50 | 16.9 |
| 9 | NEUUA147CCV | ALS | Visit 1 | 58–62 | Female | Limb | 45 | 0.12 | 85.5 |
| 14 | NEUKV735AUM | ALS | Visit 1 | 58–62 | Female | Limb | 27 | 1.75 | 52.3 |
| 16 | NEUHC282LHU | ALS | Visit 1 | 53–57 | Female | — | 36 | 2.00 | 43.6 |
| 17 | NEUBD395VAU | ALS | Visit 1 | 63–67 | Female | Limb | 29 | 0.71 | 36.1 |
| 18 | NEUWD980ECC | ALS | Visit 1 | 68–72 | Male | Limb | 44 | 0.67 | 28.8 |
| 19 | NEUZM834HCJ | ALS | Visit 1 | 63–67 | Male | Limb | 38 | 1.67 | 23.6 |
| 20 | NEUDL746XER | ALS | Visit 1 | 63–67 | Female | Limb | 40 | 0.22 | 37.3 |
| 23 | NEUEG527ZFL | ALS | Visit 1 | 63–67 | Male | Limb | 30 | 3.00 | 43.1 |
| 25 | NEUVD353HEG | ALS | Visit 1 | 63–67 | Female | Limb | 39 | 0.25 | 25.0 |
| 26 | NEUWL422HH2 | ALS | Visit 1 | 58–62 | Female | Limb | 38 | 0.83 | 65.8 |
| 27 | NEUHX995ZCH | ALS | Visit 1 | 33–37 | Male | Limb | 44 | 0.33 | — |
| 32 | NEUUR827CBY | ALS | Visit 1 | 68–72 | Female | Limb | 31 | 1.42 | 88.4 |
| 33 | NEUDA898CDV | ALS | Visit 1 | 63–67 | Female | Limb | 38 | 1.67 | 33.3 |
| 34 | NEUEJ732BTX | ALS | Visit 1 | 63–67 | Male | Limb | 45 | 0.50 | 56.5 |
| 4 | NEUYN637ZLE | Control | Visit 1 | 23–27 | Male | — | — | — | — |
| 7 | NEUAU836MWR | Control | Visit 1 | 18–22 | Male | — | — | — | — |
| 10 | NEUTK792LKM | Control | Visit 1 | 58–62 | Male | — | — | — | — |
| 11 | NEUWX096VVV | Control | Visit 1 | 53–57 | Female | — | — | — | — |
| 12 | NEUTX727BTX | Control | Visit 1 | 48–52 | Male | — | — | — | — |
| 13 | NEUYF348DNX | Control | Visit 1 | 38–42 | Female | — | — | — | — |
| 15 | NEUBE941UYL | Control | Visit 1 | 68–72 | Female | — | — | — | — |
| 21 | NEUNN201YH7 | Control | Visit 1 | 43–47 | Female | — | — | — | — |
| 22 | NEUZG882BZB | Control | Visit 1 | 68–72 | Male | — | — | — | — |
| 24 | NEUUB259TW5 | Control | Visit 1 | 43–47 | Female | — | — | — | — |
| 28 | NEUKY048BU4 | Control | Visit 1 | 53–57 | Female | — | — | — | — |
| 29 | NEUFT927MH2 | Control | Visit 1 | 73–77 | Male | — | — | — | — |
| 30 | NEUPL452UJJ | Control | Visit 1 | 33–37 | Male | — | — | — | — |
| 31 | NEUDZ824VB2 | Control | Visit 1 | 48–52 | Female | — | — | — | — |
| 35 | NEUUL863MX6 | Control | Visit 1 | 58–62 | Female | — | — | — | — |
| 36 | NEUFB501FNY | Control | Visit 1 | 53–57 | Female | — | — | — | — |
| 37 | NEUDM239AHB | Control | Visit 1 | 43–47 | Female | — | — | — | — |
| 38 | NEUBK916UHZ | Control | Visit 1 | 33–37 | Female | — | — | — | — |
| 39 | NEUZB565VR8 | Control | Visit 1 | 43–47 | Male | — | — | — | — |
| Site of onset, ALSFRS-R total score, early slope, and NfL concentration are only collected for ALS-spectrum patients; "—" denotes not applicable or not available. | | | | | | | | | |

**Supplementary Table 1**. Cohort table with all 39 patients (20 ALS-spectrum + 19 non-neurological controls), Visit 1, with the requested columns: Code, Patient ID, Subject Group, Visit, Age, Sex, Site of Onset, ALSFRS-R Total, Early Slope (pts/mo), NfL (pg/mL)

| Covariate | Imbalanced vs. group? | Drives the AE signal? | Verdict |
|---|---|---|---|
| **Age** | Yes (p=0.026, ALS older) | No — within-group partial ρ=−0.12 (p=0.46); age not decodable from the latent ($R^2$<0); retraining on an age-matched cohort reproduces the separation | Not a confounder |
| **Sex** | No (p=0.75) | No | Not a confounder |
| **Ethnicity** | No (p=0.23) | No | Not a confounder |
| **Race** | No (p=0.49, ~all White) | No variation | Not a confounder |
| **Clinical-trial coenrollment** | Yes (p=$1.4\times10^{-4}$) | No — within-ALS p=0.49–0.82 | Collinear with disease; no within-group effect |
| **Riluzole** | ALS-only | No — within-ALS p=0.55–0.75 | Signal is not a drug effect |
| **Edaravone** | ALS-only | No — within-ALS p=0.88–1.00 | Signal is not a drug effect |
| **El Escorial severity** | ALS-only | No — p=0.49–0.81 | Not severity-driven |
| **ALSFRS-R total** | ALS-only | No — ρ=−0.04 to −0.13 | Not severity-driven |
| **SVC (% predicted)** | ALS-only | No — ρ=−0.08 to −0.19 | Not respiratory-driven |
| **Disease duration** | ALS-only | No — ρ=+0.08 to +0.12 | Not duration-driven |
| **Onset site** | 18 limb / 1 mixed | — | No variation |
| **Batch / acquisition brightness** | not recorded | No — global EEM intensity AUC=0.52 (p=0.83) vs. disease AUC=0.99 | Signal is spectral shape, not intensity |

**Reference model performance**
dim477 ALS/control AUC = 0.987 • P_ALS out-of-fold AUC = 0.961

**Supplementary Table 2. Sensitivity analyses for confounding**. We evaluated every covariate recorded for the cohort against both model readouts (the aggregated latent feature dim477 and the out-of-fold classifier probability), reasoning that a variable can only bias the case/control signal if it is both imbalanced between groups and associated with the model output. Age was the only demographic difference (ALS 58.0 ± 10.8 vs. control 49.2 ± 14.3 years, $p = 0.026$) but did not confound the signature: the latent feature was flat with age within each group (partial $\rho = -0.12$, $p = 0.46$), age was not recoverable from the representation (within-group cross-validated $R^2 < 0$, versus AUC = 0.955 for group), discrimination was unchanged in a 1:1 age-matched subset (medians 55 vs. 56 years, $p = 0.91$; AUC = 0.986), and retraining the entire model on that age-balanced cohort reproduced the separation (out-of-fold AUC = 0.78), a value indistinguishable from size-matched but age-imbalanced subsets (0.78, range 0.72–0.85), confirming the modest reduction reflected the smaller training set rather than removal of the age imbalance. Sex, ethnicity, and race were balanced between groups (all $p \geq 0.23$) and unrelated to the signal. Disease-modifying therapy is collinear with diagnosis and therefore cannot be fully separated from disease, but within the ALS arm the signature did not differ between patients taking versus not taking riluzole ($p \geq 0.55$) or edaravone ($p \geq 0.88$), nor did it vary with clinical-trial co-enrollment, El Escorial category, ALSFRS-R score, respiratory function, disease duration, or genetic subtype (all n.s.), indicating a disease-state marker rather than a treatment or severity readout. Finally, although acquisition order was not logged, per-spectrum normalization removes global intensity differences and gross fluorescence intensity did not discriminate the groups (AUC = 0.52, $p = 0.83$), arguing that the signature reflects within-spectrum shape integrated across the three incubation timepoints rather than a batch or brightness artifact
